# Defining the disturbance in cortical glutamate and GABA function in psychosis and its origins and consequences

**DOI:** 10.1101/2024.06.26.24308831

**Authors:** Bill Deakin, Elizabeth Liddle, Mohanbabu Rathnaiah, Cathy Gregory, Mohammad Katshu, Gemma Williams, Silke Conen, Richard Smallman, Loes C. Koelewijn, Adriana Anton, Jyothika Kumar, Lauren E. Gasgoyne, Chen Chen, Naghmeh Nikkheslat, John Evans, Bernard Lanz, James Walters, Peter Talbot, Lena Palaniyappan, Krish D. Singh, Peter Morris, Steven R. Williams, Peter F. Liddle

**Affiliations:** Division of Psychology and Mental Health, University of Manchester, Manchester Academic Health Sciences Centre M13 9PT, UK; Institute of Mental Health, Division of Mental Health and Clinical Neuroscience, University of Nottingham, Nottingham, NG7 2TU, UK; Nottinghamshire Healthcare NHS Foundation Trust, Duncan McMillan House, Nottingham, UK. Medical; CUBRIC, School of Psychology, College of Biomedical and Life Sciences, Cardiff University, CF24 4HQ, UK; Division of Medical Education, University of Manchester, M19 3PT; Sir Peter Mansfield Imaging Centre, University of Nottingham, Nottingham NG7 2RD, UK; Stress, Psychiatry and Immunology Lab & Perinatal Psychiatry, The Maurice Wohl Clinical Neuroscience Institute, King’s College London UK; CIBM Center for Biomedical Imaging, Lausanne, Switzerland. Animal Imaging and Technology, EPFL, Lausanne, Switzerland; MRC Centre for Neuropsychiatric Genetics and Genomics, Cardiff University, CF24 4HQ, United Kingdom; Robarts Research Institute, Western University, London, Ontario, Canada; Department of Medical Biophysics, Western University, London, Ontario, Canada; Department of Psychiatry, Western University, London, Ontario, Canada; Lawson Health Research Institute, London, Ontario, Canada; Douglas Mental Health University Institute, Department of Psychiatry, McGill University, Montreal, Quebec, Canada; Division of Informatics, Imaging & Data Sciences, University of Manchester, Manchester M13 PT, UK

**Author notes:** Joint senior authors (Steven R. Williams, Peter F. Liddle).

## Abstract

It is widely thought that the onset of psychotic symptoms in schizophrenia may arise from an early neurotoxic phase, possibly related to oxidative stress or inflammation, and a late residual damage phase associated with persistent negative symptoms. We tested this hypothesis in a 3-centre study using magnetic resonance spectroscopy (MRS) to determine whether abnormalities in glutamate, glutamine and GABA content in anterior cingulate cortex (ACC) differed between people with minimally treated ‘Recent’ onset schizophrenia and an ‘Established’ group with > 10 years of treatment. We tested whether neurochemical abnormalities were i) mediated by raised circulating inflammatory cytokine concentrations, c-reactive protein (CRP) and interleukin-6 (IL-6), or depletion of glutathione and ii) associated with ratings of positive and negative symptoms. Relative to age-matched controls, the Established group showed significantly greater reduction in ACC glutamate than the Recent group, which did not differ from controls. This effect was not attributable to antipsychotic drug exposure. Patient ACC glutathione was negatively correlated with age. IL-6 was increased in both clinical groups, while increases in CRP were greater in the Established than Recent group. Elevated CRP was entirely accounted for by greater antipsychotic drug exposure and BMI, while residual elevation in IL-6 in the Established group did not account for their lower ACC glutamate. GABA was reduced relative to controls across ACC and occipital voxels. This reduction was not associated with drug treatment, BMI or cytokine levels. Only ACC GABA content correlated significantly with symptoms, lower content with greater positive and negative symptoms across both groups.

## Introduction

Interest in the role of cortical glutamate and GABA neurochemistry in the pathogenesis of schizophrenia dates back almost half a century [1]. This has been greatly reinforced in the last decade by genome wide association studies (GWAS) which have increasingly revealed that many risk genes cluster around the pre and post-synaptic elements of glutamate and GABA synapses [2–4]. Imbalance between excitatory glutamate and inhibitory GABA function has been implicated in schizophrenia and neurodevelopmental disorders and in processes such as neurotoxicity and neuro-inflammation that influence onset of symptoms, treatment response and long-term outcome [5–8]. The onset of psychosis has long been attributed to an exaggeration or neurotoxic pathology of neurodevelopmental cortical synaptic pruning which is most active in late adolescence [9–11]. Indeed it is increasingly clear from longitudinal imaging studies that loss of frontal cortical grey matter occurs close to the time of transition from the prodrome to psychosis, possibly within a few weeks[12].This may involve loss of synapses rather than loss of neurons as has been reported in histological and gene-expression studies in post mortem brain [13, 14]. Here we investigate the role of abnormal cortical glutamate and GABA synaptic function in the clinical pathogenesis of early and established stages of psychosis and whether inflammation and oxidative stress are mediating mechanisms.

Olney and Farber proposed that impaired NMDA-glutamate receptor signalling is a core deficit in schizophrenia that causes dysfunctional glutamate release[15]. The psychotomimetic effects of drugs that impair NMDA neurotransmission (phencyclidine or PCP, and ketamine) appear to be mediated by disinhibition of glutamate release through failure of restraint by inhibitory GABA interneurons [16, 17]. GABA feedback inhibition is driven by NMDA glutamate receptors on GABA interneurons. In rodent models of psychosis, PCP-like drugs ultimately induce regionally specific excitotoxic damage to glutamate neurones, loss of parvalbumin containing GABA interneurons and cognitive impairment [18, 19]. In humans also, post-mortem brain studies consistently report impaired function in parvalbumin and somatostatin containing subtypes of GABA interneurons [20, 21]. Equally, GABA changes could be the primary driver of glutamate changes and are compatible with the long-standing GABA deficiency theory of schizophrenia[22]. The role of GABA and whether disinhibition of glutamate release occurs early in psychosis and eventual synaptic loss underpins chronicity and negative symptoms are key issues for the present study.

There have been many magnetic resonance spectroscopy (MRS) studies of glutamate and GABA content in brain regions in people with schizophrenia. In keeping with the NMDA model, a landmark meta-analysis by Marsman et al reported overall increased MRS glutamine in medial frontal cortex (MFC)/anterior cingulate cortex (ACC) but decreased glutamate in cohorts comparing patients versus controls and that both metabolites decreased with age of the cohorts at faster rate than controls [23]. The extent to which these static measures reflect glutamate release rather than neuronal integrity, synaptic content and turnover is debatable. However, since synaptic glutamate is converted to glutamine by astrocytes, increases in the latter may have been a better index of increased excitatory neurotransmission in early psychosis, albeit less accurately measured at 3T, than glutamate itself.

There are obvious confounds in combining different cohorts and centres, studied at different MR field strengths and there have been no explicit comparisons of MRS glutamate in early vs established psychosis in the same study. Furthermore, effects of age are confounded with cumulative drug exposure. Indeed, a recent mega-analysis reported decrements in MFC/ACC glutamate in proportion to age and to daily antipsychotic drug exposure[24]. We sought to detect evidence of glutamate/glutamine disinhibition in early phase psychosis and loss in the established phase and to identify potential origins in drug treatment, oxidative stress or neuroinflammation, and the consequences for clinical pathogenesis.

The discovery that rare autoimmune disorders with antibodies against NMDA receptors can present as schizophrenia was a remarkable corroboration of the NMDA deficiency theory while also reinforcing interest in the neuroinflammatory hypothesis of schizophrenia [25]. Circulating cytokine concentrations such as Interleukin-6 (IL-6)-6 and C-reactive protein (CRF) are consistently increased across many cytokine screening studies in schizophrenia [26]. Whether peripheral cytokines penetrate the brain to induce a damaging microglial neuroinflammatory state and grey matter loss [27] or exert direct effects on synaptic development, function and survival is debated [28–30]. We tested these ideas by measuring peripheral cytokines and 2 putative indices of central inflammation, MRS glutathione content in ACC and in one centre, PET radioligand binding to the translocator protein (TSPO), a putative marker inflamed microglia [31]. We examined whether the central or peripheral markers mediated or influenced the group differences in MRS glutamate and GABA measures or their association with clinical symptoms.

We aimed to test the over-arching hypothesis that symptoms of schizophrenia arise from phase-dependent dysfunction in cortical glutamate/GABA synapses; an early neurotoxic phase, possibly related to oxidative stress or inflammation, and a late phase of residual damage, the phases relating respectively to acute psychosis and persistence of negative symptoms. We recruited matched samples of Recent-onset patients with non-affective psychosis with less than 12 weeks of treatment and a group with more than 10 years of treatment together with age-appropriate controls across 3 research centres. The variables of primary interest were the 4 MRS metabolites, glutamate, glutamine, glutathione, and GABA in ACC. The same measures in occipital cortex (OC) were part of a separate connectivity workstream which also enabled us to replicate a previous report of low OC GABA and test for a transcortical deficit [32]. We tested pre-specified predictions using analysis of variance to contrast early and late phase differences between controls and patients. We used correlation and mediation analysis to test for the influence of medication-related confounds, mechanistic inflammatory markers and ROS on the 4 MRS variables and their association with positive and negative symptoms and cognitive performance.

## Methods

The study received ethical approval from the National Research Ethics Service Committee Northwest – Lancaster UK, reference 14/NW/0298 on 18/06/2014.

### Participants

Participants with schizophrenia under the care of local psychiatric services were recruited by clinical research associates who prepared a case-summary for a consensus diagnosis process between 3 clinicians [33]. All patient participants currently met DSM IV criteria for schizophrenia, schizoaffective disorder, or schizophreniform disorder. Two samples of patients (aged 18–55 years and fluent in English) were recruited at each site, from populations representing two phases of illness: a minimally treated group (less than 12 weeks) with recent onset of illness of less than 5 years (Recent group) and a group with established illness of at least 10 years duration (Established group). Two groups of healthy Control participants were recruited locally by public advertisement and selected to match the patient groups (site-wise) for age, sex, and parental occupation using the National Statistics Socio-economic classification (NS-SEC), 5-class self-coded method [21]. The intended final sample size was 60 patients (20 per site) for each Phase Group, and 30 healthy controls (10 per site) age matched to each of the patient groups giving a planned total sample of 180. For full inclusion and exclusion criteria see Supplementary Section 1.

### Clinical variables

We recorded years of education and body-mass index (BMI) for all participants. Additional measures for patients included duration of illness; current antipsychotic defined daily dose (AP-day); and a 11-point rating of lifetime antipsychotic exposure measure (AP-life) reflecting both dose and duration of medication with fixed criteria for each level (Supplementary Material Section 2). Symptom measures included Positive and Negative symptom scores assessed using PANSS [34]. Cognitive function in all participants was assessed using a short-form of the Wechsler Adult Intelligence Scale, 3^rd^ Ed. [35] WAIS-III [36] validated for schizophrenia [37], and consisting of four subtests: Digit-symbol coding; Information; Block Design; and Arithmetic [37]. Subtest scores were combined to give a Full Scale IQ Standard Score (FSIQ). We used the Wechsler Test of Adult Reading (WTAR) as a proxy for pre-morbid IQ [38]. Psychosocial functioning was assessed using the Personal and Social Performance Scale (PSP) [39].

### MRS spectroscopy

Metabolites of interest were glutamate, glutamine, glutathione, and GABA, and were measured in two voxels, one placed bilaterally in the anterior cingulate cortex (ACC, 35x40x20 mm^3^) and one in occipital cortex (OC, 30x30x30 mm^3^at 3T, 28x28x28 mm^3^at 7T). 3T scanners were used at Cardiff and Manchester and a 7T scanner at Nottingham. Short-echo spectra (PRESS TE/TR =35/2000ms at 3T, STEAM TE/TM/TR =17/17/2000 ms at 7T) were acquired at all sites. At 7T the STEAM spectra were used to measure all 4 metabolites, while at 3T MEGA-PRESS was used to measure GABA (TE/TR = 68/2000 ms) and glutathione (TE/TR = 130/2000). Unsuppressed water acquisitions were acquired from each voxel to provide a reference for absolute quantification and corrected for voxel CSF (Supplementary Section 3). For full MRS details for each site see Supplementary MRS Methods. MRS measures were referenced to water, consistent with best practice as described in the recent consensus articles on acquisition and processing of human brain 1H MRS data [40, 41]. See Supplementary Section 4 for site normalisation and extreme values. See Supplementary Table 3 for Descriptive statistics for raw MRS and cytokine values by site and group.

### Cytokines

Venous blood samples were collected and centrifuged within one hour at 1300–2000g for 10 min, and plasma stored at -70°C degrees at each site prior to shipping in batches for assay. Cytokines were measured using Meso Scale Discovery (MSD) V-plex immunoassays (human) kits (MSD, Rockville, USA) [42, 43]. Any cytokine value below the standard Minimum Detectable Value (MDV) was replaced with the MDV. We log_10_-transformed the positively skewed assayed values, then site-normalised them (as for the MRS values) to control for small but systematic site differences.

### Statistical Design, hypothesis testing, and analysis

The study was designed to have 80% power at an alpha of .05 to find differences in ACC glutamate abnormality between phases (Phase x Diagnosis interaction). We selected ACC glutamate as the primary synaptic measure being more accurately determined than glutamine at 3T. The expected effect size was based on Marsman et al.’s meta-analysis showing increasing deficits in ACC glutamate with mean sample age with a slope of -0.04±0.03 per year of age [23]. This indicates a medium effect size (Cohen’s d =0.6) for comparisons between groups differing by 15 years. As the primary analysis, uncorrected *p* values are presented for this test. We predicted that low GABA in patients would be evident regardless of phase of illness and in either or both, ACC and OC voxels and therefore tested for main effects of Diagnosis in ACC, OC, and the voxel mean. For ACC glutathione, we predicted depletion in early or both phases, tested respectively by Diagnosis x Phase interaction and a main effect of Diagnosis. The analysis plan is summarised in Supplementary Section 7. We applied an FDR adjusted alpha correction for these five additional tests, and report both uncorrected *p* values and FDR corrected *q* values (Supplementary Tables 6 and 7).

For all other hypothesis tests, including planned follow-up tests of effects of Diagnosis within each Phase group, and a minimal set of mechanistic correlational predictions specified in the proposal (listed unchanged below), we present uncorrected *p* values as measures of model fit [44, 45] taking *p*<.05 as ‘evidence in support of the hypothesis’, and *p*>.1 as ‘no evidence to support the hypothesis’.

- Reduced GABA content will be associated with increased ACC glutamate or glutamine or both in Recent onset cases.
- Positive symptoms will be associated with greater ACC glutamate and reduced GABA in Recent onset patients whereas negative symptoms will be associated with low ACC glutamate in established patients.
- In early schizophrenia, depleted ACC glutathione in Recent onset patients will be associated with increased ACC Glu and increased circulating CRP and IL-6 concentrations.

### Statistical methods

We used ‘robust’ methods for all descriptive and inferential statistics, implemented in R version 4.1.2 *Rcpre team* [46]using functions from the WRS2 package (Supplementary Section 5) [47]. Robust methods are a family of methods that are robust to deviations from the distributional assumptions of parametric methods (e.g. long-tailed distributions), and to excess leverage from outlying values. For mediation models we used the lavaan structural equation modelling (SEM) package, version 0.68 [48], with the MLR estimator (Maximum Likelihood estimation with Robust standard errors) and removed outlying values prior to analysis. Variables with skewed distributions were transformed to symmetrical distributions (log_10_ for BMI and cytokine values; square root for AP-day).

## Results

### Participant characteristics

There were 62 (15 female) patients and 35 (9 female) controls in the Recent group and 76 (23 female) patients and 39 (14 female) controls in the Established group, slightly exceeding the target sample sizes (Supplementary Table2). Summary statistics and group comparisons for demographic, cognitive, and clinical variables are shown in Table 1. Details on missing or unusable data and are given in Supplementary Tables 3 and 4.

**Table 1.** Demographic, cognitive and clinical variables by group. Legend. Tr-Diff is difference between 20% trimmed means. P values are for Yuen’s robust t-test for differences between trimmed means. NS-SEC p values for the linear-by-linear association χ^2^ test). P values <.05 are shown in bold. WTAR and FSIQ values are Standard Scores (pop. Mean=100, SD=15), PSP - Personal and Social Performance, a control -patient difference, ^b^Recent -Established difference, ^c^p value computed using log_10_-transformed scores

|  | Recent |  |  |  | Established |  |  |  | Patients |  |
| --- | --- | --- | --- | --- | --- | --- | --- | --- | --- | --- |
|  | Controls | Patients | Difference<sup>a</sup> |  | Controls | Patients | Difference<sup>a</sup> |  | Difference<sup>b</sup> |  |
| Demographic | Mean (SD) |  | Tr-Diff. | P | Mean (SD) |  | Tr-Diff. | P | Tr-Diff. | P |
| Age | 24.4 (4.4) | 24 (5.1) | -0.60 | .521 | 42.3 (7.8) | 42.4 (7.5) | 0.28 | .883 | 19.6 | N/A |
| Parental class (NS-SEC) | 2.7 (1.7) | 2.9 (1.6) | N/A | .635 | 2.9 (1.6) | 2.8 (1.8) | N/A | .721 | N/A | .750 |
| Years of education | 16.5 (2.6) | 14.7 (2.3) | -2.16 | **.001** | 17 (3.6) | 14.5 (3.1) | -2.81 | **.001** | -0.5 | .377 |
| BMI | 24.1 (3.6) | 25 (4.4) | 0.01 | .988 | 25.8 (4) | 29.8 (5.6) | 4.40 | **.000** | 5.4 | **.001** |
| Current IQ (FSIQ) | 108.9 (22.5) | 94.3 (17) | -18.75 | **.000** | 105.2 (16.2) | 89.9 (14.2) | -15.17 | **.000** | -3.5 | .293 |
| Pre-morbid IQ (WTAR) | 106.8 (17.5) | 100.3 (18.6) | -8.43 | **.025** | 108.4 (9.7) | 100 (17.1) | -7.13 | **.017** | -0.1 | .982 |
| FSIQ-WTAR | -2.1 (14.9) | 5.9 (15.3) | -8.83 | **.002** | 3.1 (14.2) | 10.1 (14.8) | -7.88 | **.011** | -5.2 | .054 |
| **Clinical** |  |  |  |  |  |  |  |  |  |  |
| Antipsychotic score |  |  |  |  |  |  |  |  |  |  |
| AP-day (0-4) |  | 0.7 (0.7) |  |  |  | 1.3 (0.8) |  |  | 0.7 | **.000** |
| AP-life (0-10) |  | 1.7 (0.8) |  |  |  | 8 (1.3) |  |  | 6.2 | **.000** |
| PANSS Negative |  | 14.5 (6.0) |  |  |  | 13.4 (5.3) |  |  |  | .495<sup>c</sup> |
| PANSS Positive |  | 12.5 (5.2) |  |  |  | 13.2 (6.1) |  |  |  | .286<sup>c</sup> |
| PSP |  | 63.5 (14.2) |  |  |  | 56.4 (13.5) |  |  | -8.1 | **.011** |

Mean scores on all cognitive measures were lower in the patient groups than in their age-matched control groups and they had fewer years of education. Decline from premorbid IQ (FSIQ-WTAR difference) was greater for both patient groups than their controls. The Recent and Established patient groups did not differ appreciably on years of education nor cognitive test scores, but the Established group had a lower mean PSP score. Decline from premorbid IQ (FSIQ-WTAR) was slightly greater in the Established Group. Patient groups did not differ appreciably in severity of Positive and Negative symptoms. Daily antipsychotic dose and mean cumulative antipsychotic exposure were both greater in the Established group. Mean BMI was also greater in the Established than in the Recent patients, as well as higher than their age-matched control group.

### Glutamate and GABA: effects of diagnosis and phase

Greater reductions in ACC glutamate were found in Established patients than in the Recent group relative to their age-matched controls, as predicted, and evidenced by a statistically significant Diagnosis x Phase interaction, Q=5.74, N=183, *p=*.022 (Figure 1A). There were no significant interactions with Site. Follow-up tests found no evidence of elevated glutamate concentration in the Recent onset group (ES = 0.13 (95% CI: -0.32, 0.59), *p=*.574 but there was large effect size for the reduction in ACC glutamate in the Established group relative to controls, ES = -0.59 (95% CI: -1.14, 0.09), *p=*.012. ACC glutamine showed no evidence of a Diagnosis x Phase interaction (Figure 1B), Q=0.60, N=180, *p=*.440.

**Figure 1.**
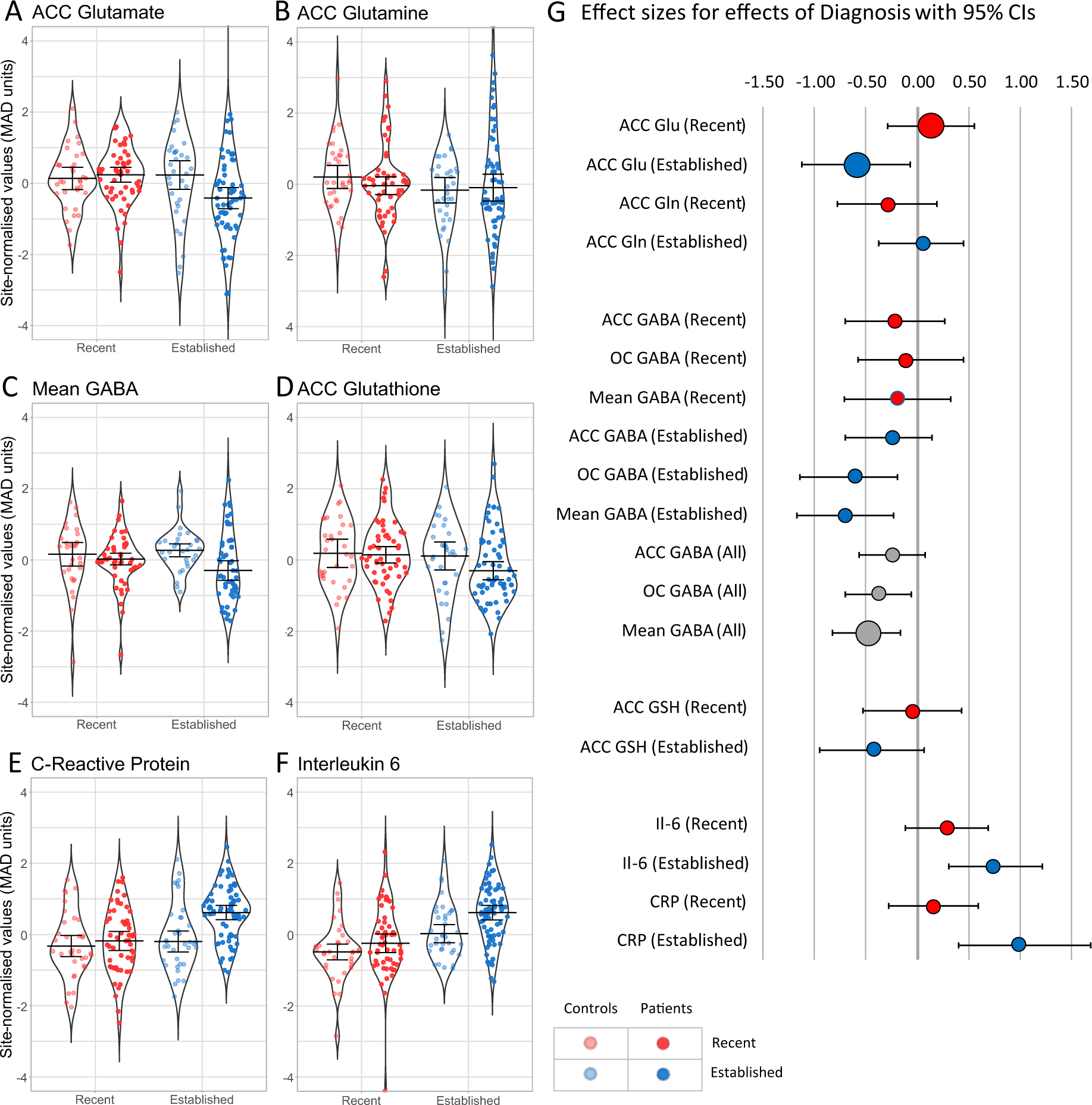
Effects of Diagnosis and Phase on MRS variables and cytokines. Effects of Diagnosis for MRS variables (A to D) and inflammatory cytokines (E and F). Panel F shows robust effect sizes (AKP, a robust version of Cohen’s d) for effects of diagnosis, with 95% CIs. Supplementary Table 5 shows robust effect sizes and CIs for effect of diagnosis on all variables in each group.

In keeping with a generalised GABA deficit in psychosis, average GABA concentrations (ACC & OC) were lower in patients (Figure 1C), Q=8.08, N=155, *q=*.036, *p=*.006), with a Medium ES = -0.48 (95% CI: -0.83, -0.16) for the effect of Diagnosis (Figure 1G). There was no evidence for interactions between Diagnosis and either Phase (*p=*.102), or Voxel (*p=*.999, as tested by a 3-way ANOVA with ACC-OC difference as the dependent variable).

### Glutathione and Cytokines

ACC glutathione was not depleted relative to controls overall (*q*=.296, *p=*.197), nor was there any Phase x Diagnosis interaction (*q*=.350, *p=*.175). CRP and IL-6 concentrations (Figure 1E and F, ST3) showed a similar pattern of greater elevation relative to controls in Established than in Recent onset groups. However, the Diagnosis x Phase interaction was significant only for CRP (*p=*.047; IL-6 *p=*.159) whereas both showed main effects of diagnosis (CRP *p=*.004; IL-6 *p=*.003). In patients, both cytokines were highly correlated with each other across phase groups (*r=*0.50, *p*<.001 Figure 3).

### Correlation analysis of mechanistic markers and confounders

To test the specified mechanistic correlates of glutamate and GABA dysfunction, robust correlations were computed between the 4 ACC MRS variables and potential influencing variables within groups and, using group mean-centred data, across both patient groups. They are summarised in Figure 2 and shown in detailed scatter plots and matrices in Supplementary Figure 1 and Table 8).

**Figure 2.**
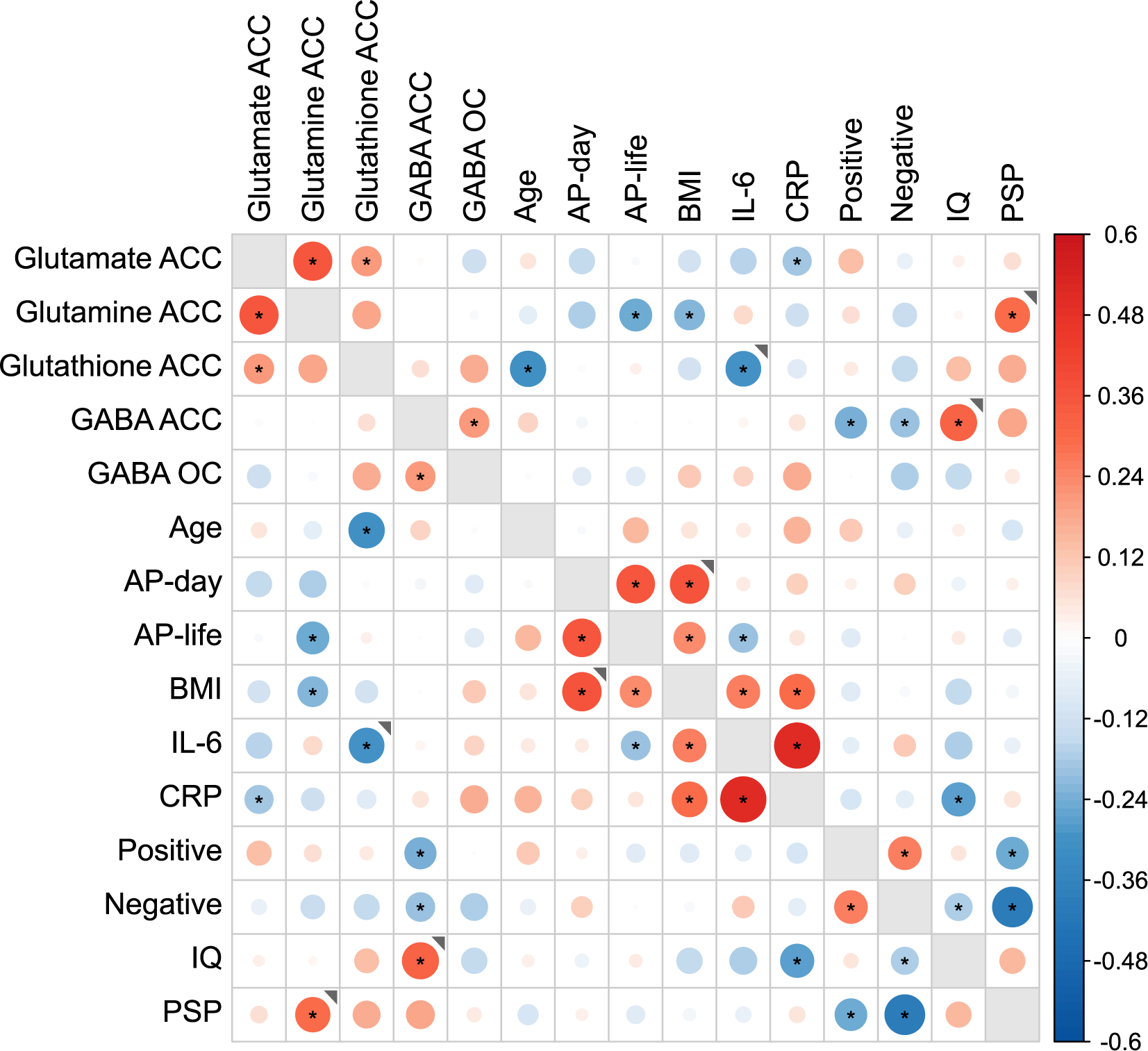
Correlation matrix for MRS variables, potential mediators, and clinical correlates. Correlation matrix relating 4 MRS dependent variables (top left) for all patients (n=>100) to potential mediators and to clinical correlates. Size and colour intensity indicate magnitude of correlation, red for positive and blue for negative correlations scaled from -0.6 to + 0.6 using group mean centred data. Dots indicates all p values <.05. Correlations are across both groups except where the grey marker ◥ indicates that the correlations differed between groups at p<.01 and was p<.05 only in the Recent onset group. There were no instances where the correlation was greater and p<.05 only in the Established group. Where groups correlations differed but both at p<.05 in the same direction, the overall average is shown.

Glutamate, glutamine and glutathione content were positively intercorrelated (Figure 2 top left) but none correlated with GABA. There was no evidence of the predicted inverse relationship between decreased GABA content and increased glutamate or glutamine content. Glutamate did not show any significant correlations with clinical measures predicted or otherwise. Glutamine inversely correlated with drug exposure and BMI. In contrast, decreased GABA content correlated only with clinical measures: lower values correlated with greater PANSS positive and negative symptoms, across both groups (PANSS positive: *r*=-.24, *p*=.014; PANSS negative: *r*=-.20, *p*=.040), with lower IQ, in the Recent onset group, *r=*0.36, *p*=.025), and poorer social functioning (PSP) across both groups, *r=*0.19, *p=*0.051).

In terms of early inflammatory mechanisms, although glutathione was not depleted in the Recent group, the predicted inverse association with increased levels of IL-6 was observed within this group (*r=*-.30, *p=*.037). Only glutathione correlated with age, inversely and within and across each phase group. CRP and IL-6 levels were inversely correlated with ACC glutamate across both groups, significantly so for CRP, suggesting a possible mediating mechanism for low ACC glutamate in Established patients. Neither daily nor lifetime drug exposure was associated with lower ACC glutamate or raised CRP and IL6 levels (middle of fig2). However, greater lifetime exposure to antipsychotic drugs was associated with higher BMI which in turn showed its well-known association with increased cytokines (CRP and IL-6). Nevertheless, lifetime drug exposure was associated with lower IL-6 concentrations (*r=*-.20, *p*.023), a possible anti-inflammatory effect which might mask illness-related increases in IL6 that could mediate the changes in ACC glutamate.

### Mediation analysis of reduced ACC glutamate in Established patients

The correlation analyses (Figure 2) identified several possible influences that might partially or wholly account for the greater ACC glutamate deficit in Established patients. These included effects related to AP medication exposure, but also potential mediation by co-occurring phase differences in circulating inflammatory cytokines (CRPι1 and IL-6ι1). We evaluated these possibilities in a sequence of mediation models in which we expressed the values of MRS and inflammatory variables as deviations from their age-matched control means (adjusted variables denoted by ι1).

There was no evidence that the negative Phase effect on glutamateΔ (deficit greater in the Established group) was mediated by daily and/or lifetime antipsychotic dose (Figure 3A); none of the indirect effect pathways reached *p*<.1 and the direct (unmediated) effect of Phase on ACC glutamateΔ remained evident (green line, Figure 3A), *β* =-0.57, *p*<.023. However, as the correlation analyses indicated a negative correlation between AP-life and ACC glutamine, we also investigated mediation via the effects of AP-life on ACC glutamineΔ (Supplementary Figure 3B). We found weak evidence (*p*<.1) for partial mediation of the negative Phase effect on ACC glutamateΔ by AP-life via a negative effect on ACC glutamineΔ (*β* =-0.18, *p=*.066). However, this effect was offset by an indirect positive effect of Phase on ACC glutamineΔ itself (*β*=0.19, *p=*.059), and accounting for these indirect effects strengthened the evidence for the direct effect of Phase on ACC glutamateΔ (*β*=-0.66, *p=*.002)

**Figure 3.**
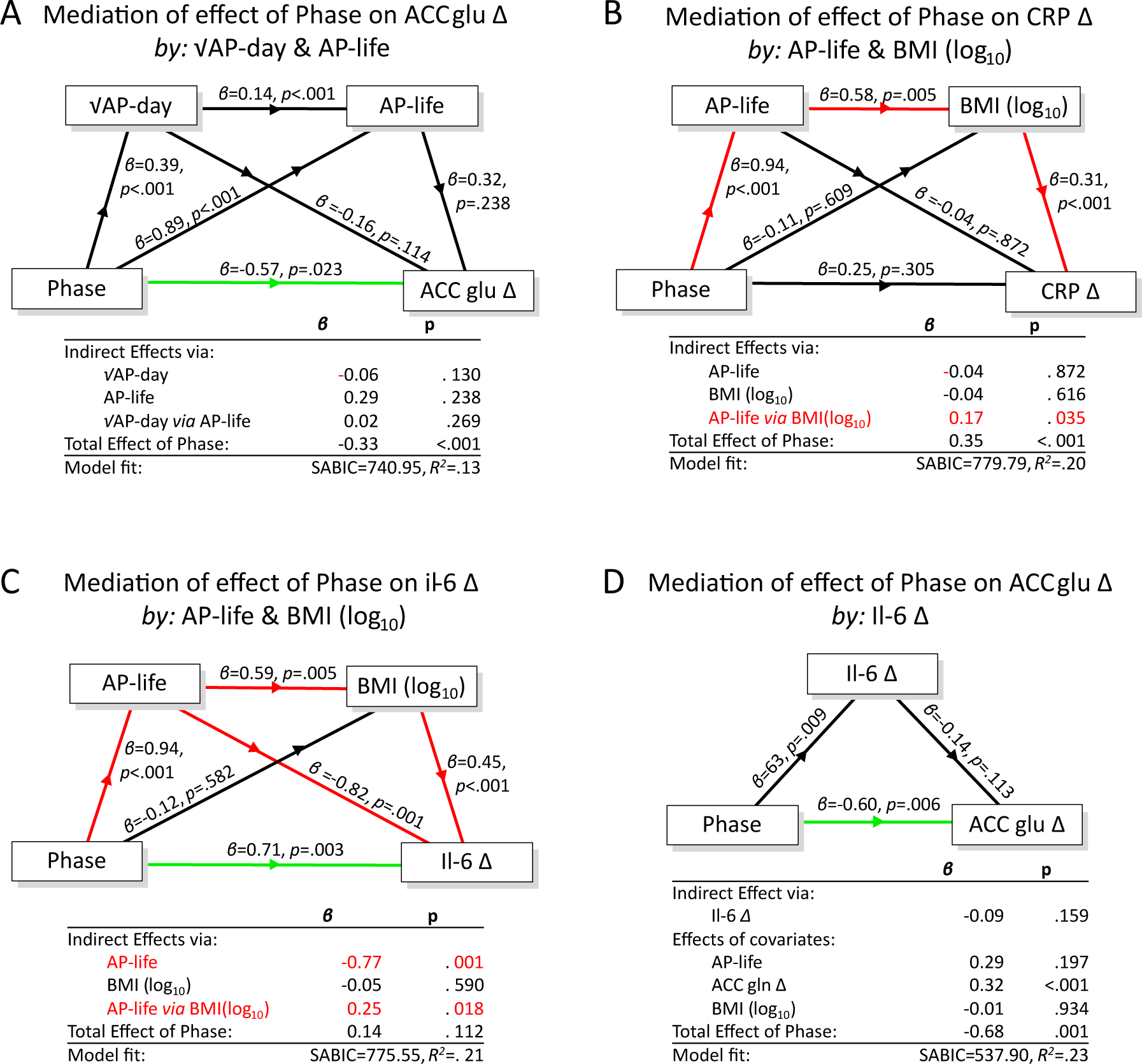
Mediation analysis of phase-dependent loss of ACC glutamate. Models A to C: medication-related mediators of Phase effects on ACC glutamate, CRP and IL-6. Model C revealed excess IL-6 in the Established group (p-003 for the Direct effect of Phase) after accounting for medication-related effects. Model D: Phase-related increases in IL-6 did not mediate the phase-related reductions in ACC glutamateΔ (p=.159 for the Indirect Effect). Coefficients are standardised β values. Total Effect = sum of direct and indirect effect of Phase. Indirect pathways are shown in red if p<.05. Direct non-mediated effect of Phase shown by horizontal connector, green if p<.05. Note that in Model D, the Total Effect excludes indirect effects mediated by the covariates.

The total Phase effect on CRPΔ (greater in the Established group; p<.001) was completely mediated by the indirect pathway from Phase via AP-life and BMI (Figure 3B and Supplementary Figure 3C) leaving no direct i.e. unmediated effect (p>.1). However, for IL-6Δ, while AP-life acted via BMI to increase IL-6Δ, this was offset by the unexpected negative effect of AP-life on IL-6Δ itself, as noted in the correlation analysis, which was independent of BMI (Figure 3C red diagonal). Accounting for these opposed indirect effects revealed evidence for an underlying direct effect of Phase on IL-6Δ (*β* =0.71, *p=*.003), namely a residual excess of IL-6Δ in the established group over levels in the Recent onset group, possibly indicative of a Phase-dependent effects of illness.

We investigated whether the revealed effect of Phase on IL-6Δ levels might mediate the ACC glutamateΔ Phase-effect. The Phase effect on IL6 was isolated (Figure 3D, β =.63, *p=*.009) by covarying the opposing indirect effects of AP-life and BMI shown in Figure 3C. By including glutamineΔ in the covariates, we controlled for its previously mentioned indirect influences on the effect of Phase on glutamateΔ. This model provided no evidence to support an indirect (mediating) effect of excess IL-6Δ on Phase-associated ACC glutamateΔ (Figure 3D, *p=*.159) and the direct effect of Phase remained evident (β= -0.60, *p=*.006). See Supplementary Section 9 for a full account of the mediation model series).

## Discussion

This study tested the hypothesis that excessive glutamate neurotransmission in early psychosis underpins the emergence of positive symptoms and results in loss of synaptic glutamate and the development of persistent negative symptoms in established illness. We confirmed with well-matched samples recruited explicitly from patients with early and established illness, that the Established group had greater ACC glutamate deficits but found no evidence of excess in the Recent group. The greater deficit in the Established group was not attributable to greater drug exposure in the Established group and was not associated with greater negative symptom scores. There was a generalised decrease of GABA content across ACC and OC which did not correlate with glutamate changes but was associated with greater positive and negative symptoms across both groups. We found no evidence of ACC glutathione abnormalities in patients, although some evidence that might indicate a greater depletion with age. In inverse contrast to ACC glutamate, circulating cytokine concentrations of CRP and IL-6 were greater (significantly for CRP) in established illness than controls raising the possibility of a causal inflammatory process.

In the Recent onset patients, there was no increase in glutamate or glutamine in ACC and neither correlated with positive symptoms as we had predicted. Thus, we found no evidence that dysregulation of glutamate occurs in early psychosis or underlies symptoms. Indeed, early reports of increased glutamine in early psychosis [23] have not been replicated in more sensitive studies at 7T [49–52] with a single report of a trend-level association with symptoms. It is possible that the MRS measures are too static or diffuse in origin to detect dysfunctional glutamate release in early schizophrenia. It is also possible but perhaps unlikely that a neurotoxic process underlying the onset of symptoms had resolved so rapidly as to be undetectable by group differences in MRS measures or a greater spread than controls given that half the Recent group were within 7 months of onset. Nevertheless, the existence of a small neurotoxic subset could be hidden in the variability as suggested by some within-patient studies [53, 54].

There was no evidence of ongoing oxidative stress in Recent onset patients; there was no depletion of glutathione nor were levels of circulating cytokines CRP markedly increased as might have been expected had there been inflammatory neurotoxicity undetected by MRS ACC glutamate or glutamine. However, il-6 was elevated relative to controls across groups and ACC glutathione was negatively correlated with il-6 (Figure 2) in the Recent group, a possible hint of early inflammatory central oxidative stress. As previously reported in the Manchester subset, there was no evidence of central microglial inflammation in Recent onset patients as ACC TSPO radioligand binding was not increased in Recent or Established patients [31]. In summary, there was no MRS evidence of abnormal ACC glutamate, glutamine or glutathione content in Recent onset minimally treated patients with schizophrenia and minimal evidence of evidence of systemic or central immune activation.

The decreases in glutamate in the Established group are broadly in keeping with published studies and meta-analyses. In a meta-analysis of 65 studies in medial prefrontal cortex including ACC, Merritt et al reported reductions in patients with schizophrenia and no changes in glutamine[55]. Some meta-analyses report more marked decrements in ACC glutamate in older cohorts as first reported by Marsman et al [23]. In a mega-analysis combining individual data, both daily antipsychotic drug exposure and age were associated with greater reduction in ACC glutamate [24]. However, our Defined Daily Dose measure showed only a minimal inverse association with ACC glutamate only in the Established group (*r*=-0.24, *p=*0.06; Supplementary Table 8), and this did not mediate the phase-related reduction in ACC glutamate (Supplementary Figure 9B). Additionally, we observed for the first time that long-term cumulative exposure to antipsychotic drugs had no effect on ACC glutamate (Figure 3A). However, such exposure was associated with reduced ACC glutamine across both phase groups and so an influence of antipsychotic drug exposure on synaptic glutamate function cannot be ruled out.

Reduced ACC glutamate in the Established but not the Recent group is compatible with the proposed accumulation of neurotoxic damage to glutamate synapses in established schizophrenia [15, 56]. This is corroborated by recent PET studies in which the synaptic marker SVA2 was reduced in patients with established illness but not first episode patients [57, 58]. Furthermore, SVA2 correlated with MRS ACC glutamate content in healthy controls and in patients but to a lesser extent because of their constrained range of reduced levels of SV2A binding. The duration of illness in our Established group spans 25 years but duration was not associated with greater loss of ACC glutamate suggesting that the loss is static after 10 years of treatment or stably maintained by an active process. The contribution to pathogenesis is unclear since reduced glutamate did not show the predicted correlation with negative symptoms (table 2) nor was this observed in a metanalysis of 15 studies with age-related decrease in MRS creatine-scaled glutamate [24]. Indeed, the meta-analysis found that against the background of reduced levels, glutamate reliably but weakly correlated with positive symptoms as we also observed (Established *r=*.21; *p=*.111).

We evaluated the possibility that previous or current inflammation contributed to the static reduction in ACC glutamate. Circulating CRP and IL-6 concentrations were increased in the Established group and were weakly predictive of reduced ACC glutamate (Figure 2). However, mediation analysis indicated that whereas that late phase increases in CRP were accounted for by associated treatment and BMI differences, the effect of Phase on IL-6 was enhanced after adjustment for treatment confounds. Nevertheless, there was no evidence that IL-6 increases directly mediated the late phase decrease in ACC glutamate (Figure 3D). Indeed, low ACC glutamate was recently associated with polygenic risk for reduced NMDA receptor function shared with treatment resistance albeit in a younger cohort than the Established group [59].

Reduced ACC glutamate correlated at p<.05 with lower glutathione (Figure 2) and although glutathione levels were not reduced in patients, it alone among MRS and cytokine variables, correlated inversely with age within each phase group. This might suggest that excessive levels of ageing-associated reactive oxygen species indexed in the brain by the age-MRS glutathione decline could induce detectable damage to glutamate terminals in Established which are not detectable in Recent onset schizophrenia in keeping with a recent meta-analysis [60]. In the periphery, reactive oxygen species could underly the chronic mild inflammation indexed by increased IL-6 concentrations associated with schizophrenia [61, 62].

There is consistent evidence for impaired GABA synthesis in cortical interneurons in samples of post-mortem brain in schizophrenia, but it is less clear whether GABA neurones are lost [20]. The interpretation of In-vivo MRS GABA studies is complicated by variable definitions of ROIs and by clinical and treatment status. In 17 MRS studies with ROIs ranging from subgenual, pregenual, anterior, and dorsal cingulate, Egerton et al (2017) found no overall decrease in GABA [63]. More recently, Simmonite et al reported an overall decrease in GABA in 10 studies in ROIs corresponding to our ACC ROI [64]. The authors suggest GABA deficits may be modulated by illness stage (greater in acute) or ameliorated by prolonged medication. However, their subgroup difference was small and we found no evidence for an effect of phase or of medication exposure on GABA deficits in patients. Iwata et al (2018) found 3 studies in OC all 3 reporting reductions in GABA in keeping with our study [65]. Our MRS findings are strongly corroborated by a report of lower GABA concentrations measured directly in cerebro-spinal fluid in 40 prospectively recruited patients with FEP and 21 controls [66]. Furthermore, reduced CSF GABA correlated with PANSS positive scores and functional outcomes as in our study in which positive and negative symptoms correlated with low GABA across Recent and Established groups. The findings suggest that low GABA may have a key role in the pathogenesis of symptoms in schizophrenia but not apparently acting via glutamate disinhibition.

### Limitations

This is a cross-Sectional study and therefore we cannot conclude that differences between recent onset and established cases arise from longitudinal change within cases. It is possible that there was an accumulation of cases with more persistent or severe illness, associated deficits in MRS metabolites and higher cytokine levels in the Established group. Although the overall clinical sample is large, the Recent and Established groups were designed to have half the number of controls (n=30) as patients (n=60) to increase the power of within-patient analyses but at the cost of power for control-patient comparisons. The multi-site design will have introduced noise owing to differing clinical and healthy populations sampled. Furthermore, although MRS procedures and analyses were co-ordinated, they were not completely harmonised in order to take advantage of local expertise and manpower, as detailed in Supplementary material. This may have contributed to site effects in metabolite quantification, not completely controlled for by statistical procedures explained in Supplementary Material.

## Conclusions

In testing the neurotoxic synaptic loss account of schizophrenia pathogenesis, we confirmed the prediction that MRS glutamate in anterior cingulate cortex is reduced in people with long-term treated symptoms of schizophrenia. The deficit appears to be the stable result of a process which is not apparent in early-stage illness, is not related to exposure to chronic antipsychotic medication and is not mediated by central or peripheral measures of inflammation. There was no evidence of the predicted association with negative symptoms or with other symptoms. GABA content was reduced in anterior cingulate and occipital cortex in the full sample of patients with recent or established schizophrenia. Reductions in anterior cingulate GABA content correlated with positive and negative symptoms, but did not correlate with markers of inflammation, degree of drug treatment or with glutamate. These in-vivo findings strengthen the evidence that impaired GABA function is involved the pathogenesis of the symptoms of schizophrenia. Further analyses to be reported elsewhere are relating these findings to cortical connectivity using magnetoencephalography and assessing dynamic glutamine-glutamate turnover using ^13C^-MRS.

## Supporting information

Supplementary MRS Methods

## Acknowledgements

This project was funded by Medical Research Council Experimental Medicine Challenge Full Grant (MR/K020803/ 1). LP acknowledges research support from Monique H. Bourgeois Chair in Developmental Disorders and Graham Boeckh Foundation (Douglas Research Centre, McGill University) and a salary award from the Fonds de recherche du Quebec-Sante’ (FRQS).

## Data availability

The database for this study database will be made available on reasonable request to

