## Supplementary MRS Methods for "Defining the disturbance in cortical glutamate and GABA function in psychosis and its origins and consequences"

**Acquisition details**

Acquisition details for each site are contained in the following Tables, which encompass the information suggested in the consensus article on reporting of MRS studies (1):

- Table S1 Scanners and Coils
- Table S2 Sequence Parameters and Processing Software
- Table S3 Voxel sizes and locations
- Table S4 Acquisition parameters
- Table S5 MEGAPRESS acquisition details
- Table S6 $T_1$-weighted anatomical image acquisitions

*Table S1: Scanners and Coils*

| Site | Scanner | Field | Receive Coil |
|---|---|---|---|
| Cardiff | GE HDx | 3.0T | 8 channel head coil |
| Manchester | Philips Achieva | 3.0T | 32 channel head coil |
| Nottingham | Philips Achieva | 7.0T | 32 channel head coil |

*Table S2: Sequence Parameters and Processing Software*

| Site | Sequence | TE/(TM)/TR ms | Aves | Processing |
|---|---|---|---|---|
| Cardiff / Manchester | PRESS[a] | 35/2000 | 128 | jMRUIv6 QUASARY |
| Cardiff / Manchester* | GABA MEGAPRESS[b] | 68/2000 | 300/296* | GANNET 2.0 |
| Cardiff / Manchester | GSH MEGAPRESS[c] | 130/2000 | 256 | jMRUIv6 AMARES |
| Nottingham | STEAM[d] | 17/(17)/2000 | 288 | LCModel |

*Citations: a(2); b(3, 4);c(5);d(6)*

*Table S3: Voxel Sizes and Locations*

| Site | Voxel Location | Voxel Size mm |
|---|---|---|
| Cardiff / Manchester / Nottingham | Bilateral ACC | 35x40x20 |
| Cardiff / Manchester / Nottingham* | Occipital Cortex | 30x30x30 / 28x28x28* |

*Table S4: Acquisition details*

| Site | Sequence | BW (Hz) | pts |
|---|---|---|---|
| Cardiff | PRESS | 2000 | 1024 |
| Cardiff | MEGAPRESS | 5000 | 4096 |
| Manchester | PRESS / MEGAPRESS | 2000 | 1024 |
| Nottingham | STEAM | 4000 | 4096 |

*Table S5: MEGAPRESS Acquisition (Cardiff, Manchester)*

| Metabolite | TE (ms) | Frequency offsets (edit/control) | Edit pulse shape | Length/BW |
|---|---|---|---|---|
| Glutathione | 130 | 4.56/1.44 ppm | gauss | 14ms/106Hz |
| GABA | 68 | 1.90/7.6 ppm | gauss | 14ms/106Hz |

*Table S6: $T_1$-weighted anatomical images*

| | Sequence | TI/TE/TR (ms) | Matrix | Voxel size |
|---|---|---|---|---|
| Cardiff | FSPGR* | 450/88/3.5 | 256x256x192 | 1mm isotropic |
| Manchester | IR-TFE* | 1150/3.9/8.4 | 256x256x90 | 0.9x0.9x1.9mm |
| Nottingham | IR-TFE* | 987/3.4/7.3 | 256x256x180 | 1mm isotropic |

*3D Fast Spoiled Gradient-Recalled acquisition; Inversion Recovery prepared Turbo (rapid) Field Echo (MPRAGE (7))

**Processing of Spectra**

Spectroscopic analysis was divided among the 3 centres in order to make best use of the human resources provided by the funding agency and to take advantage of local expertize, such as Cardiff's experience in measuring GABA at 3T, Manchester's experience in processing short-echo PRESS and glutathione MEGAPRESS at 3T and the expertize in Nottingham in 7T MRS. Three processing packages were used: GANNET (8, 9); jMRUI (10-13); LCModel (14). Thus MEGAPRESS GABA data were quantified in Cardiff using GANNET (15), PRESS TE35 and glutathione MEGAPRESS were quantified in Manchester using jMRUIv6 (QUASARY and AMARES respectively(16)) and in Nottingham spectra were quantified using LCModel (17). Each centre used established protocols as detailed further in the cited articles.

$T_1$-weighted anatomical images were used for accurate voxel placement and image segmentation (Table S6, Figure S1). Cardiff and Manchester images were segmented in Cardiff using GANNET 2.0, while Nottingham images were segmented in Nottingham using SPM8, in both cases based upon Ashburner's and Friston's method (18).

**Water referencing**

Non water-suppressed acquisitions were acquired in each centre. Using the tissue water signal as reference, molar metabolite concentrations were calculated after correction for the CSF content of the voxel. In Nottingham a tissue specific correction of the water signal was used following Gasparovic et al (19), whereas in Cardiff and Manchester the 'standard' correction employed by LCModel was used which does not account for differences in grey matter and white matter within the spectroscopic voxel. While this will affect absolute values, as the tissue content was not significantly different across groups or centres, comparisons of concentration remain valid.

*Fig. S1 Voxels in ACC and OC overlaid in yellow on anatomical $T_1$-weighted images. Subject was chosen from Manchester.*

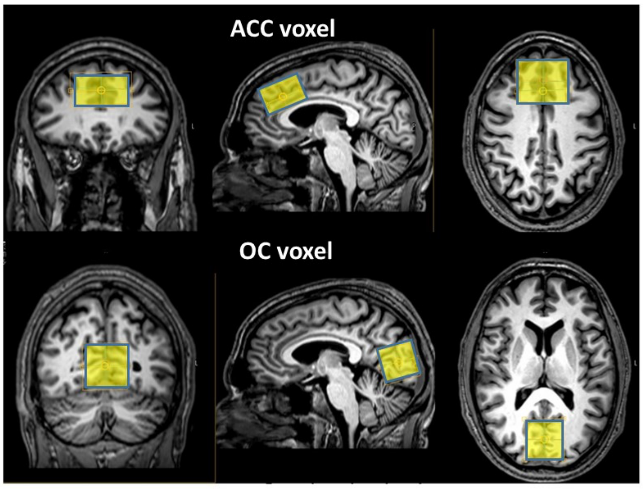


**Quality of Spectra**

Figs. S2 and S3 show all the short echo spectra that were analyzed from each centre aligned and overlaid on one plot for each voxel location.

*Fig S2/S3 Overlay of spectra from ACC and OC. All spectra were loaded into jMRUI v7.0, frequency aligned, residual water removed and intensities normalized by the tissue water signal. No line-broadening was applied for display. Assignments: NAA – N-acetylaspartate, Glut – glutamate; Crn – creatine + phosphocreatine; Cho – choline-containing compounds; m-Ino – myo-inositol*

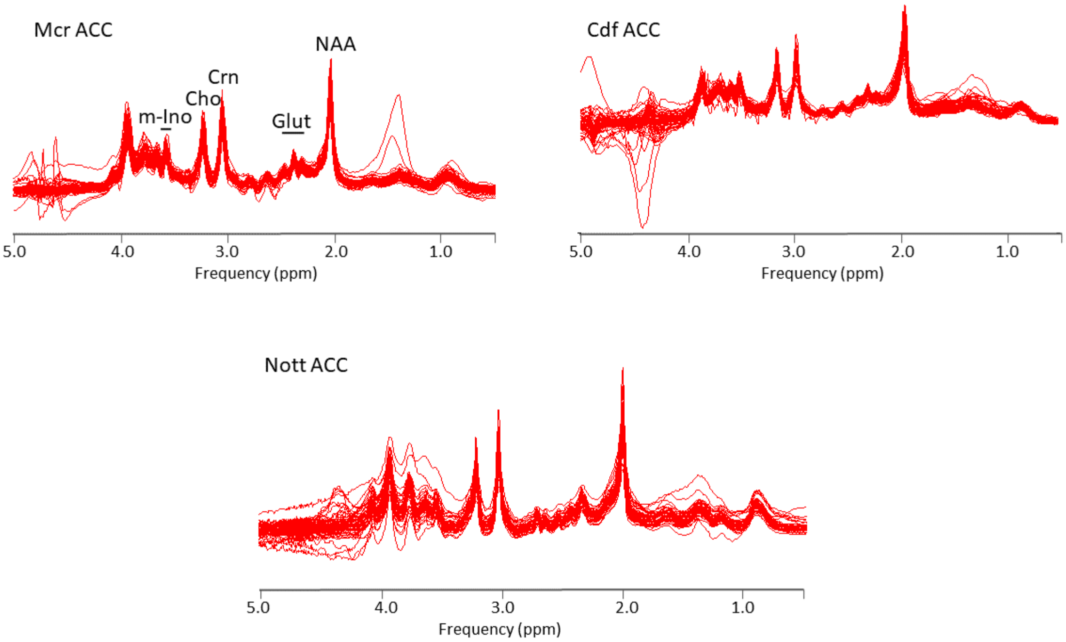

*Fig S3 Overlay of short echo spectra from OC. Processed as above.*

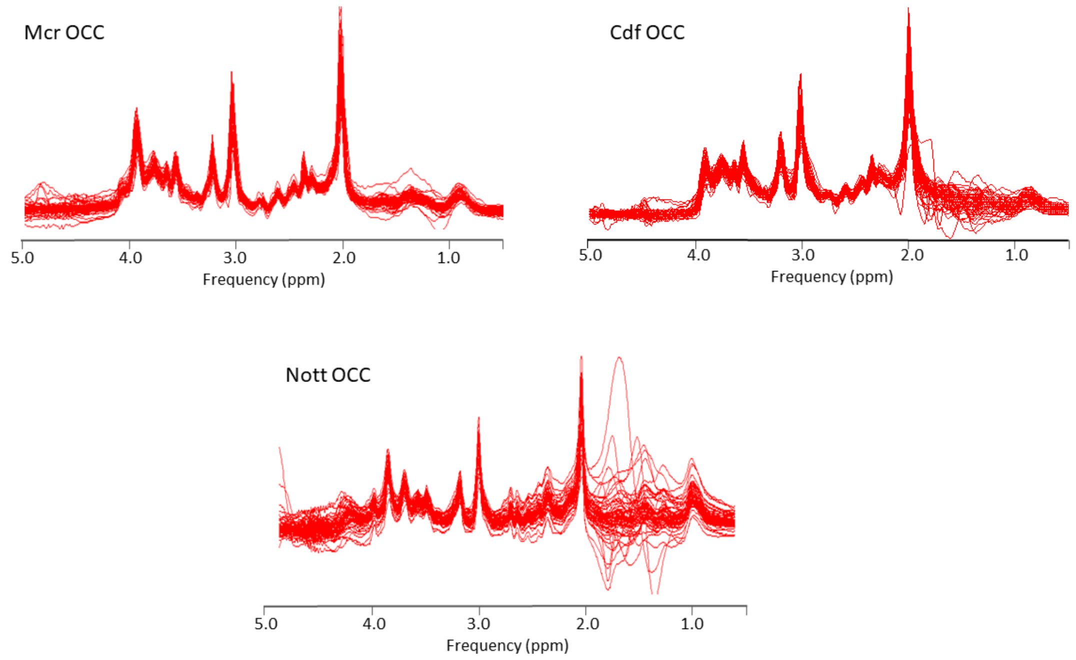


Most spectra from the ACC are free of any significant lipid contamination or obvious artefacts arising from poor water suppression, whereas in the OC the 7T spectra show a number of examples with lipid contamination, though this does not affect the spectra between 2 and 4 ppm. In general the appearance of the spectra is highly consistent with high signal-to-noise ratio and well resolved resonances.

MEGAPRESS spectra from the ACC of all analysed subjects are shown in Fig.S4 for both GABA and glutathione sequences. The relatively long echo times (68ms for GABA, 130ms for glutathione) combined with the low concentration of the metabolites mean the spectra have worse signal-to-noise than the short echo spectra, but in all cases the GABA and glutathione resonances (centred at 3.02 and 2.98 ppm respectively) are quantifiable.

*Fig.S4 Overlaid MEGAPRESS spectra from Manchester and Cardiff. Spectra were frequency aligned, line-broadened for display (6Hz) and the intensities normalized to tissue water. Assignments: NAA – N-acetylaspartate; $GABA^+$ - edited GABA plus co-edited macromolecules; GSH – edited cysteinyl residue of glutathione; (N) asp – edited aspartyl resonances of NAA, aspartate and N-acetylaspartylglutamate*

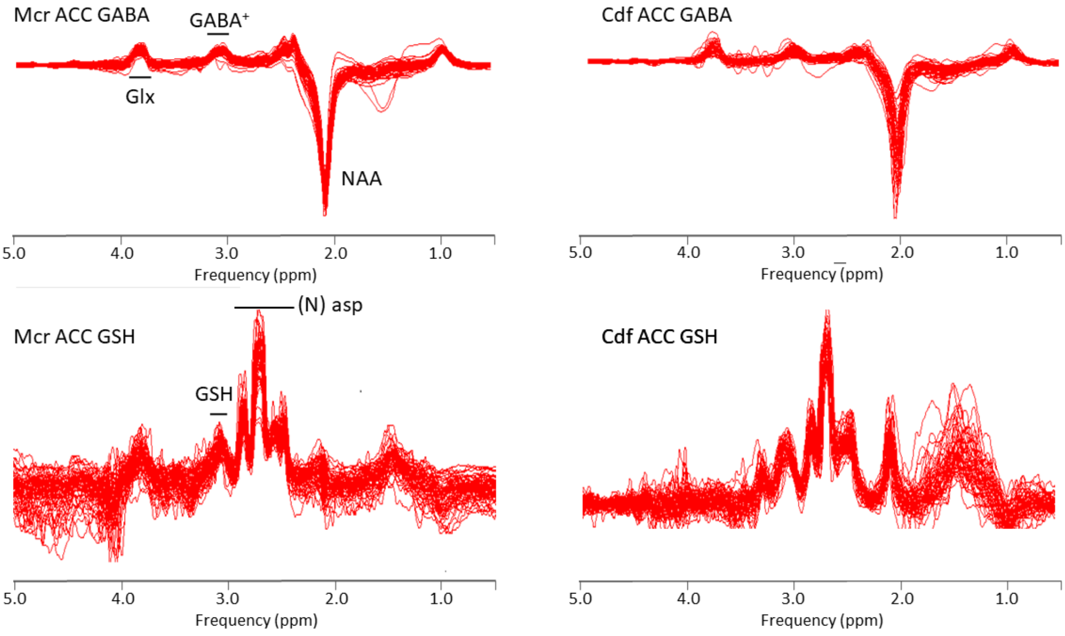

**Quantification Results**

Examples of the output of the fitting routines for the short echo spectra are shown below for data from Cardiff and Manchester (Fig.S5) and Nottingham (Fig.S6).

*Fig. S5 jMRUI fit to Cardiff and Manchester ACC spectra. QUASAR-Y was used to fit the data, with the baseline estimated using the Subtract-QUEST option. The raw data and the estimate are shown in the bottom trace; the estimate with the baseline in the second trace, with individual components and the residual in the upper two traces. A basis set of 6 metabolites and 2 macromolecule components was fitted to the data.*

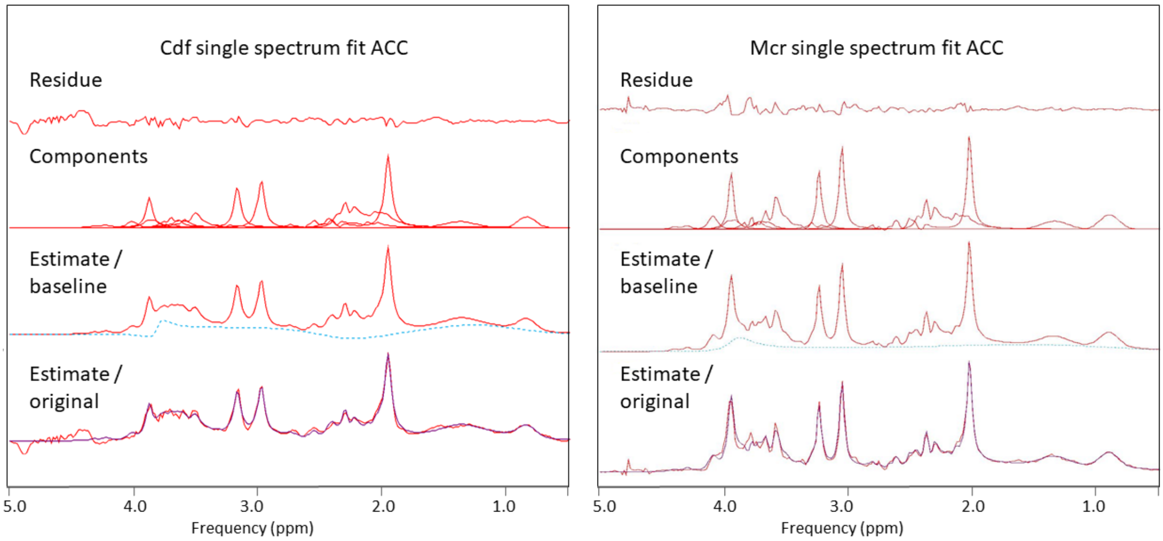

*Fig S6 LCModel output for example ACC and OC spectra from Nottingham. Individual fitted components for glutamine, glutamate, GABA and glutathione are shown in the lower traces, together with the residual (bottom) and the original data overlaid on the fit (top). 20 metabolite components plus a macromolecular baseline were fitted to the data.*

**Nott LC Model Fits**

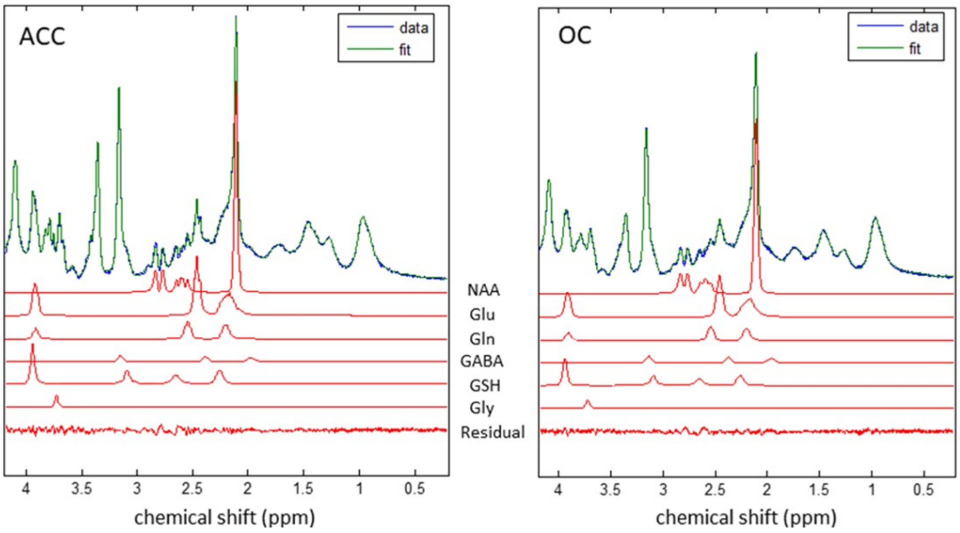


The edited 3T MEGAPRESS spectra were quantified using GANNET in Cardiff for the GABA data and in Manchester using jMRUIv6 for glutathione. Examples of the output of the fitting routines are shown in Figs.S7 (glutathione/AMARES) and S8 (GABA/GANNET).

*Fig.S7 AMARES output from glutathione analysis. The glutathione cysteine resonance centred at 2.98 ppm was fitted as a doublet following Kaiser et al. (20), the aspartyl resonances of N-acetyl aspartate were also modelled to improve baseline definition around 3 ppm. The estimate and original data are shown in the bottom trace with the fitted glutathione components and residual above.*

**AMARES fits to ACC glutathione spectra**

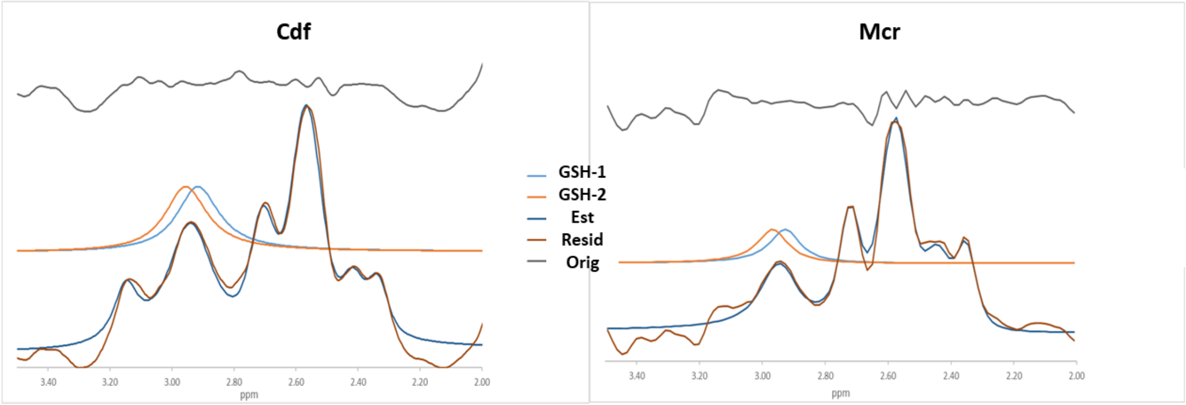

*Fig.S8. Examples of spectra fitted by GANNET from Cardiff and Manchester.*

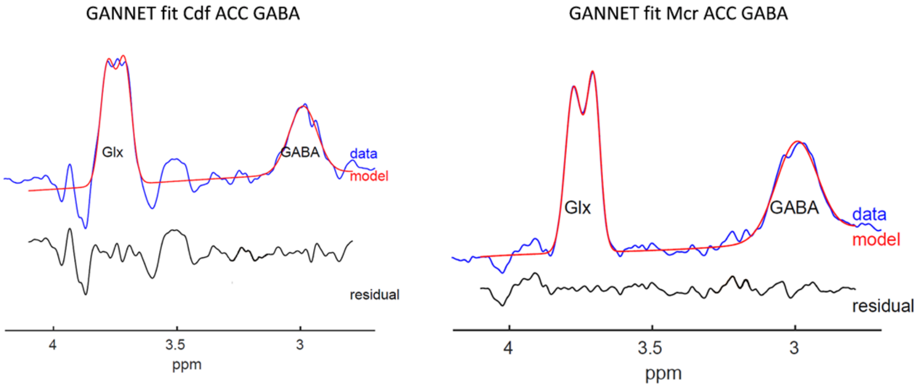


**Quality Assurance Metrics**

In accordance with recent guidelines on reporting MRS data (1) the following metrics are reported for the short echo spectra in Table S7: water and N-acetylaspartate (NAA) linewidth and %CRLB for NAA and glutamate.

*Table S7. Quality Metrics for short echo spectra*

| Site / brain region | Water LW (Hz) | | NAA LW (Hz) | | NAA %CRLB | | Glut %CRLB | |
|---|---|---|---|---|---|---|---|---|
| | ACC | OC | ACC | OC | ACC | OC | ACC | OC |
| Cardiff | 9.3±2.0 | 9.0±3.2 | 8.8±2.1 | 9.2±2.0 | 0.29±0.14% | 0.23±0.05% | 0.78±0.47% | 0.77±0.39 |
| Manchester | 6.8±0.7 | 6.9±0.5 | 9.6±1.2 | 9.7±0.9 | 0.24±0.07% | 0.22±0.001% | 0.61±0.18% | 0.80±0.30 |
| Nottingham | 12.3±1.3 | 13.6±1.9 | 15.1±3.4 | 14.9±2.2 | 1.06±0.42% | 1.94±2.32% | 1.20±0.45% | 2.34±2.20% |

Consistent with the appearance of the overlaid spectra, the average linewidths with coefficients of variation of 10-20% indicate good spectroscopic quality, consistent shimming and good reproducibility. The larger % CRLB for the Nottingham data are a consequence of using a more complex basis set with more components than were used for the jMRUI fitting for the Manchester and Cardiff data. There is a decrease in Bias and increase in Variance as more components are added into the basis set.

The fitted linewidths and % CRLB are reported below (Table S8) for GABA and glutathione measured using MEGAPRESS in Cardiff and Manchester.

*Table S8 Quality metrics for MEGAPRESS spectra.*

| Metabolite / Site | GABA | | Glutathione | |
|---|---|---|---|---|
| | LW (Hz) | % CRLB | LW (Hz) | % CRLB |
| Cardiff | 16.1±1.4 | 4.3±3.1 | 13.7±1.8 | 5.5±3.2 |
| Manchester | 20.3±2.3 | 4.8±1.3 | 9.7±2.4 | 14.4±6.4 |

The linewidth of the edited GABA signal is significantly larger than the N-acetylaspartate and water linewidths recorded in the same sessions. This is because the GABA signal includes a contribution from co-edited macromolecules (commonly referred to as GABA+) which was fit as a single Gaussian line. As expected from the signal-to-noise ratios of the two acquisitions and the greater complexity of the glutathione spectrum, GABA is fitted with better precision than glutathione.
