## Supplementary Material for "Defining the disturbance in cortical glutamate and GABA function in psychosis and its origins and consequences"

### Supplementary Materials

#### 1. Full inclusion/exclusion criteria

Inclusion criteria
1) Male or female, aged 18 - 55 years.
2) Ability to understand and willing to give written informed consent.
3) English as first language or fluent.
4) Current DSM IV criteria for schizophrenia, schizoaffective disorder, or schizophreniform disorder.
5) <5 years from onset for early psychosis group; or >10 years for established illness group.
6) Antipsychotic drugs: no exposure, discontinued or minimal (<12 weeks) exposure for early psychosis group; or for established illness group 8 weeks of stable treatment.

Exclusion criteria
1) Current use of any medication which may interfere with the study, in the opinion of the investigator (not including treatment for schizophrenia).
2) Clinically significant neurological disorder.
history of head injury with loss of consciousness > 5 min.
3) Current harmful use of, or recent dependence on, psychoactive substances (excluding nicotine).
4) Contraindications for magnetic resonance imaging (MRI) (e.g., claustrophobia, pregnancy, ferrous metal implants).
5) Taken part within the previous month as a participant in a clinical trial that involved taking a drug, being paid an inconvenience allowance, or having an invasive procedure (e.g., venepuncture > 50 ml, endoscopy).

Additional exclusion criteria for controls

1) Personal history of psychosis or a related disorder (as determined by the MINI-international neuropsychiatric interview v5.0.0 for DSM-IV (Sheehan et al., 1998).
2) current or recent (within 2 years) presence of depressive symptoms or treatment with antidepressant medication;
3) first degree relative with a history of psychosis.

Participants in Nottingham and Manchester were further excluded from participation if they had a blood-borne virus, relevant to the subsequent 13-C magnetic resonance spectroscopy (MRS) investigation not reported here. They were screened for diabetes and cardiovascular (ECG abnormalities). Manchester participants met criteria for inclusion in positron emission tomography (PET) studies.

Informed consent was obtained, and participants were paid an inconvenience allowance for their participation. All procedures were approved by the UK National Research Ethics Service.

#### 2. Current daily and life-time exposure to antipsychotics (from protocol)

Our aim is to define a scoring system for life-time exposure to antipsychotics which provide a meaningful estimate of exposure that is feasible to apply with the data available.

Evidence from PET studies (Kapur et al Am J Psych, 1999) indicates that antipsychotic effects are small at doses below 50% DA receptor occupancy, in the therapeutic range for 60-80% and are in excess of antipsychotic needs above 80%. However occupancy data is not available for all antipsychotics, and is not directly relevant to therapeutic effect for all antipsychotics (especially clozapine). Therefore, it is more practical to define broad ranges (low, medium and high, relative to the doses that have become established in clinical practice, even though these are only an approximate indicator of effects on brain chemistry or brain function (Leucht et al, Sz Bulletin 2016). WHO defines the Defined Daily Dose (DDD) as

> "The defined daily dose DDD is the assumed average maintenance dose per day for a drug used for its main indication in adults."

In the case of olanzapine, DDD is 10mg. According to the PET data reported by Kapur et al, during sustained treatment, olanzapine produces 50% occupancy at approximately 5mg/day; 70% occupancy at 10 mg/day and 80% occupancy at 20 mg/day. Thus, in the case of olanzapine, assuming that the therapeutic range is 50-80% DA occupancy, the therapeutic range is from 0.5 DDD to 2 DDD. For other antipsychotics, DDD is likely to be less closely related to DA occupancy. Typically DDD for first generation antipsychotics is associated with high occupancy due to the establishment of customary practice an era when higher doses were widely used, whereas for clozapine, DDD corresponds to a lower DA receptor occupancy reflecting the evidence that therapeutic effects occur at lower DA receptor occupancy than for other antipsychotics. Nonetheless, if we seek a definition of dose range based on customary practice, 0.5 DDD to 2 DDD is an appropriate estimate.

#### Dose ranges

- Low : dose < 0.5 DDD
- Medium: 0.5 DDD =< Daily Dose =< 2 DDD
- High: Daily dose > 2 DDD

#### Duration of exposure

With regard to the neural effects of life time exposure, there is little evidence to provide firm guidance, as studies are inconsistent, possibly reflecting variation between individuals based on predisposition and phase of illness, and within individual depending on brain region. Nonetheless, significant decreases in brain tissue volume have been reported in longitudinal studies of 3-5 years in the early phase of illness (Neeltje et al Arch Gen Psychiatry. 2011;68(9):871-880 https://doi.org/10.1001/archgenpsychiatry.2011.88; Kubota et al JAMA Psychiatry. 2015;72(8):803-812. https://doi.org/10.1001/jamapsychiatry.2015.0712.) Neeltljee et al reported excessive focal decreases in cortical thickness during the 5-year interval ranging from 0.05 to 0.19 mm (compared with a mean of 0.01 mm in healthy controls). Overall, evidence indicates that illness duration of 5 years is associated with substantial tissue loss in predisposed individuals with schizophrenia. If medication contributes an appreciable portion to this tissue loss, a time scale of 5 years is likely to be relevant to appreciable tissue loss.

#### Total exposure score

Range 0-10 based on duration and dose:

0 no antipsychotic exposure
1 Low dose exposure only (any duration)
2 Less than 1 year total exposure including some medium but no high dose exposure:
3 Less than 1 year of total exposure including some high dose exposure:
4 One to five years total exposure including medium but no high dose exposure lasting more than one month.

5 One to five years total exposure including with some high dose exposure lasting more than one month.
6 Five to 10 years total exposure including medium dose but no high dose exposure lasting more than one month.
7 Five to 10 years total exposure with some high dose exposure lasting more than one month.
8 Greater than 10 year exposure including medium but no high dose exposure lasting more than one month.
9 Greater than 10 years total exposure including with high dose exposure of between one month and 5 years in duration
10 Greater than 10 years total exposure including high dose exposure of greater than 5 years in duration

It is possible to re-code these scores in a manner that provides separate quantification for duration of exposure to medium or high dose antipsychotic medication and for the magnitude of sustained dose of antipsychotic medication. (In the following definitions, numbers in italics in brackets denote the coding of total exposure

#### Duration of exposure to medium or high dose antipsychotic medication

0 no exposure to antipsychotic medication (= *0*)
1 either exposure to antipsychotic medication for up to one year, or longer than one year if never exceeding low dose (= *1,2,3*)
2 exposure for one to five years including some exposure to medium or high doses (=*4,5*).
3 exposure for five to ten years including some exposure to medium or high doses (=*6.7*)
4 exposure for greater than 10 years including some exposure to medium or high doses (=*8,9,10*).

#### Magnitude of sustained dose of antipsychotic medication

0 no exposure to antipsychotic medication (= *0*)
1 Exposure only to low dose medication, or exposure to medium or high dose for less than one year (= *1*)
2 Exposure to medium doses during antipsychotic treatment exceeding one year but no exposure to high dose sustained for more than one month (= *2,4,6,8*)
3 Exposure to antipsychotic treatment exceeding one year, including exposure to a high dose sustained for more than one month (= *3,5,7,9*).
4 Exposure to antipsychotic treatment for more than 10 years including high dose treatment sustained for more than 5 years (= *10*).

#### Clozapine

It will also be useful to score clozapine exposure. DDD is not likely to be as meaningful, so four point scale is probably adequate to capture the relevant information

0 no evidence of exposure to clozapine
1 exposure to clozapine for less than 1 year
2 exposure to clozapine for greater than 1 year, less than 5 years
3 exposure to clozapine for greater than 5 years

#### 3. MRS method

MRS water-referenced metabolite concentrations were corrected for the proportion of brain tissue in each voxel by segmenting out cerebrospinal fluid from T1-weighted structural MRI scans. The Established group and their controls had greater proportions of CSF and reduced grey and white matter than the Recent groups in both ACC and OC voxels (main effects of phase) but there were no main effects ($p<.1$) of diagnosis or diagnosis by phase interactions.

As expected ACC creatine correlated positively (r=<.40) with ACC glutamate, glutamine and glutathione but we confirmed there were no significant or trend effect of group, phase or site on creatine concentrations in ACC or OCC nor did they correlate greater than r=.15 with any confound, cytokine or clinical variable

#### 4. Controlling for effects of site

Site-specific measurement factors included the scanner and spectroscopy protocols used at each site, potentially affecting the metabolite measures, and the blood sample preparation and storage facilities at each site, potentially affecting the cytokine measures.

##### MRS metabolite concentrations

Inspection of MRS metabolite values and cytokine measures at each site revealed a small number of “extreme” values (values exceeding the first or third quartile by > 3 times the interquartile range for that site). These were removed from further analysis. Substantial between-site differences remained that were not readily accounted for by differences in the characteristics of the participant samples at each site. We therefore site-normalised all values by subtracting the site median from each value and dividing by the median absolute deviation (MAD).

##### Cytokines

Although the same blood sample protocols were used across sites, and all samples were analysed by the same lab, sample preparation (e.g. centrifuge) and storage equipment were site specific, and there were also site-level differences in the time that elapsed between sample collection and assay. Inspection of the data revealed site-level differences unrelated to length of storage, and that values had a positively skewed distribution. We therefore $\log_{10}$-transformed the assayed values, and normalised them by subtracting the site median and dividing by the site mean.

#### 5. Robust statistical packages implemented in in R.

For ANOVAs, correlations and t-tests we used functions from the WRS2 package [1] for R [2] as given in Supplementary Table 1.

*Supplementary Table 1*

| Test | WRS function | Options |
|---|---|---|
| 3-way ANOVA | t3way | Trim = 20% |
| 2-way ANOVA | t2way | Trim = 20% |
| t-test | yuen | Trim = 20% |
| effect size for t-test | akp.effect | N bootstrap samples=1000 |
| correlations | pbcor (percentage bend correlation) | Beta=20% , N bootstrap samples=1000 |

For regression mediation models we used the lavaan structural equation modelling (SEM) package, version 0.68 [1], with the MLR estimator (Maximum Likelihood estimation with robust standard errors). Outliers (values exceeding the first or third quartile by > 1.5 times the interquartile range) were first removed, and skewed data transformed to a symmetrical distribution (see main text for details). As missing data for some participants for some variables were likely to be missing at random (MAR), we used casewise Full Information Maximum Likelihood (FIML) estimation to use all available data.

#### 6. Descriptive statistics

*Supplementary Table 2: Sample sizes for each group at each site.*

| Diagnosis | Site | Recent | Established | Total |
|---|---|---|---|---|
| Patients | Cardiff | 19 | 22 | 41 |
| | Manchester | 21 | 24 | 45 |
| | Nottingham | 22 | 30 | 52 |
| | Total | 62 | 76 | 138 |
| Controls | Cardiff | 10 | 12 | 22 |
| | Manchester | 10 | 14 | 24 |
| | Nottingham | 15 | 13 | 28 |
| | Total | 35 | 39 | 74 |

*Supplementary Table 3: : Descriptive statistics for raw MRS and cytokine values at each site for each group*

| | | | Cardiff | | | | | | Manchester | | | | | | Nottingham | | | | | | Total |
|---|---|---|---|---|---|---|---|---|---|---|---|---|---|---|---|---|---|---|---|---|---|
| PE | DIAGNOSIS | Dependent variable | N included | N extremes | Median | MAD | Mean | SD | N included | N extremes | Median | MAD | Mean | SD | N included | N extremes | Median | MAD | Mean | SD | N included |
| Established | Controls | ACC Glutamate | 12 | 0 | 8.11 | 1.22 | 7.76 | 1.41 | 11 | 0 | 12.12 | 2.56 | 12.37 | 2.42 | 9 | 0 | 13.40 | 1.10 | 13.18 | 1.11 | 32 |
| | | ACC Glutamine | 12 | 0 | 3.82 | 0.86 | 3.78 | 1.01 | 11 | 0 | 4.33 | 1.84 | 3.94 | 1.52 | 9 | 0 | 3.61 | 0.21 | 3.49 | 0.30 | 32 |
| | | ACC Glutathione | 11 | 0 | 2.77 | 0.85 | 2.68 | 0.99 | 12 | 0 | 3.30 | 0.39 | 3.30 | 0.70 | 9 | 0 | 3.43 | 0.42 | 3.29 | 0.44 | 32 |
| | | ACC Creatine | 12 | 0 | 5.01 | 0.94 | 4.83 | 0.98 | 11 | 0 | 8.11 | 0.61 | 8.19 | 1.17 | 9 | 0 | 12.55 | 0.76 | 12.43 | 0.96 | 32 |
| | | ACC GABA | 12 | 0 | 2.35 | 0.46 | 2.24 | 0.42 | 11 | 0 | 2.39 | 0.52 | 2.40 | 0.48 | 6 | 0 | 0.75 | 0.20 | 0.80 | 0.35 | 29 |
| | | ACC NAA | 12 | 0 | 8.63 | 0.93 | 8.30 | 1.44 | 11 | 0 | 10.52 | 0.49 | 10.56 | 0.83 | 9 | 0 | 14.24 | 0.49 | 14.01 | 1.05 | 32 |
| | | OC Glutamate | 10 | 0 | 6.87 | 1.13 | 7.04 | 2.34 | 11 | 0 | 10.94 | 0.73 | 11.20 | 0.81 | 10 | 0 | 13.14 | 0.79 | 13.57 | 1.09 | 31 |
| | | OC Glutamine | 10 | 0 | 1.02 | 0.81 | 1.36 | 1.33 | 11 | 0 | 3.70 | 0.79 | 3.86 | 0.94 | 10 | 0 | 3.68 | 0.33 | 3.85 | 0.67 | 31 |
| | | OC Creatine | 10 | 0 | 6.76 | 0.63 | 7.10 | 1.23 | 11 | 0 | 9.16 | 0.37 | 9.01 | 0.61 | 10 | 0 | 13.15 | 1.20 | 13.19 | 1.00 | 31 |
| | | OC Glutathione | 12 | 0 | 2.37 | 0.33 | 2.43 | 0.37 | 12 | 0 | 2.80 | 0.99 | 3.16 | 1.23 | 10 | 0 | 2.75 | 0.21 | 2.73 | 0.24 | 34 |
| | | OC GABA | 12 | 0 | 2.90 | 0.15 | 2.92 | 0.21 | 12 | 0 | 2.84 | 0.21 | 2.84 | 0.17 | 10 | 0 | 1.75 | 0.37 | 1.69 | 0.42 | 34 |
| | | OC NAA | 9 | 1 | 9.27 | 0.82 | 9.96 | 1.64 | 11 | 0 | 12.67 | 0.67 | 12.64 | 0.69 | 10 | 0 | 15.79 | 1.20 | 15.72 | 1.07 | 30 |
| | | C-Reactive Protein | 12 | 0 | 0.00 | 0.67 | 0.17 | 0.66 | 11 | 0 | -0.01 | 0.74 | 0.02 | 0.70 | 13 | 0 | -0.12 | 0.15 | -0.11 | 0.44 | 36 |
| | | Interleukin 6 | 12 | 0 | -0.34 | 0.16 | -0.26 | 0.26 | 11 | 0 | -0.21 | 0.18 | -0.24 | 0.20 | 13 | 0 | -0.41 | 0.09 | -0.34 | 0.21 | 36 |
| | Patients | ACC Glutamate | 19 | 0 | 6.73 | 2.52 | 6.64 | 2.09 | 23 | 1 | 11.82 | 1.41 | 12.24 | 2.38 | 21 | 1 | 12.64 | 0.99 | 12.61 | 1.01 | 63 |
| | | ACC Glutamine | 18 | 1 | 3.76 | 1.75 | 4.15 | 1.62 | 22 | 0 | 4.11 | 1.54 | 4.68 | 2.16 | 21 | 1 | 3.31 | 0.42 | 3.49 | 0.54 | 61 |
| | | ACC Glutathione | 20 | 0 | 2.52 | 0.42 | 2.87 | 0.77 | 21 | 0 | 2.38 | 0.77 | 2.67 | 0.88 | 21 | 1 | 3.21 | 0.42 | 3.24 | 0.46 | 62 |
| | | ACC Creatine | 19 | 0 | 5.01 | 0.89 | 4.99 | 1.14 | 22 | 2 | 8.25 | 0.74 | 8.19 | 0.72 | 21 | 1 | 12.63 | 0.93 | 12.61 | 0.87 | 62 |
| | | ACC GABA | 19 | 1 | 2.01 | 0.53 | 2.23 | 0.97 | 22 | 1 | 2.21 | 0.50 | 2.12 | 0.48 | 19 | 0 | 0.82 | 0.52 | 0.79 | 0.48 | 60 |
| | | ACC NAA | 19 | 0 | 7.88 | 1.87 | 8.16 | 1.75 | 24 | 0 | 11.32 | 0.70 | 11.15 | 0.72 | 20 | 2 | 13.89 | 0.52 | 13.65 | 0.73 | 63 |
| | | OC Glutamate | 18 | 0 | 6.79 | 1.39 | 6.86 | 1.23 | 23 | 0 | 11.49 | 1.08 | 11.46 | 1.16 | 16 | 0 | 13.27 | 0.89 | 13.23 | 1.51 | 57 |
| | | OC Glutamine | 18 | 0 | 0.63 | 0.88 | 0.98 | 1.16 | 23 | 0 | 4.19 | 0.77 | 4.32 | 0.95 | 15 | 1 | 3.55 | 0.43 | 3.57 | 0.50 | 56 |
| | | OC Creatine | 18 | 0 | 6.06 | 0.72 | 6.15 | 0.72 | 23 | 0 | 8.63 | 0.65 | 8.74 | 0.62 | 16 | 0 | 13.67 | 0.74 | 13.92 | 1.78 | 57 |
| | | OC Glutathione | 17 | 0 | 2.36 | 0.28 | 2.29 | 0.37 | 23 | 0 | 3.18 | 1.25 | 3.00 | 1.03 | 14 | 2 | 2.87 | 0.24 | 2.89 | 0.26 | 54 |
| | | OC GABA | 20 | 0 | 2.57 | 0.36 | 2.62 | 0.39 | 23 | 0 | 2.66 | 0.40 | 2.69 | 0.38 | 15 | 0 | 1.53 | 0.53 | 1.61 | 0.58 | 58 |
| | | OC NAA | 18 | 0 | 9.13 | 1.30 | 9.12 | 1.28 | 23 | 0 | 12.43 | 0.43 | 12.46 | 0.64 | 16 | 0 | 15.33 | 1.58 | 15.46 | 2.18 | 57 |
| | | C-Reactive Protein | 22 | 0 | 0.66 | 0.68 | 0.58 | 0.55 | 21 | 0 | 0.41 | 0.67 | 0.27 | 0.50 | 29 | 0 | 0.44 | 0.27 | 0.45 | 0.40 | 72 |
| | | Interleukin 6 | 22 | 0 | -0.06 | 0.28 | -0.06 | 0.29 | 21 | 0 | -0.23 | 0.26 | -0.24 | 0.25 | 29 | 0 | -0.11 | 0.23 | -0.12 | 0.19 | 72 |

| | | | | | | | | | | | | | | | | | | | | | |
|---|---|---|---|---|---|---|---|---|---|---|---|---|---|---|---|---|---|---|---|---|---|
| Recent | Controls | ACC Glutamate | 10 | 0 | 7.29 | 0.86 | 7.32 | 0.97 | 10 | 0 | 12.40 | 0.87 | 12.48 | 1.44 | 14 | 0 | 13.46 | 0.56 | 13.42 | 0.86 | 34 |
| | | ACC Glutamine | 10 | 0 | 4.55 | 1.09 | 4.52 | 0.81 | 10 | 0 | 4.64 | 0.99 | 4.81 | 1.40 | 14 | 0 | 3.52 | 0.39 | 3.52 | 0.43 | 34 |
| | | ACC Glutathione | 9 | 0 | 3.41 | 0.56 | 3.28 | 0.52 | 10 | 0 | 2.61 | 0.85 | 2.93 | 0.96 | 14 | 0 | 3.37 | 0.51 | 3.36 | 0.41 | 33 |
| | | ACC Creatine | 10 | 0 | 4.77 | 0.56 | 4.79 | 0.72 | 10 | 0 | 7.65 | 0.24 | 7.90 | 0.67 | 14 | 0 | 12.26 | 0.47 | 12.26 | 0.81 | 34 |
| | | ACC GABA | 9 | 0 | 2.16 | 0.90 | 2.14 | 0.99 | 10 | 0 | 2.11 | 0.59 | 2.15 | 0.58 | 13 | 0 | 0.76 | 0.18 | 0.79 | 0.27 | 32 |
| | | ACC NAA | 10 | 0 | 8.01 | 0.91 | 7.78 | 1.02 | 10 | 0 | 10.66 | 0.14 | 10.95 | 0.66 | 14 | 0 | 14.25 | 0.85 | 14.35 | 0.77 | 34 |
| | | OC Glutamate | 9 | 0 | 6.67 | 2.25 | 7.32 | 2.89 | 10 | 0 | 11.07 | 2.28 | 10.93 | 1.89 | 11 | 0 | 13.79 | 1.13 | 13.82 | 1.29 | 30 |
| | | OC Glutamine | 9 | 0 | 1.35 | 1.90 | 1.83 | 1.88 | 9 | 0 | 3.34 | 1.47 | 3.08 | 1.51 | 11 | 0 | 3.47 | 0.58 | 3.48 | 0.72 | 29 |
| | | OC Creatine | 8 | 1 | 5.89 | 0.67 | 5.87 | 0.53 | 10 | 0 | 8.71 | 0.61 | 8.62 | 0.66 | 11 | 0 | 12.72 | 0.86 | 12.85 | 1.12 | 29 |
| | | OC Glutathione | 9 | 0 | 2.69 | 0.25 | 2.65 | 0.34 | 10 | 0 | 4.00 | 0.86 | 3.80 | 1.13 | 11 | 0 | 2.77 | 0.25 | 2.73 | 0.47 | 30 |
| | | OC GABA | 10 | 0 | 2.85 | 0.33 | 2.86 | 0.28 | 10 | 0 | 2.77 | 0.30 | 2.72 | 0.44 | 11 | 0 | 1.48 | 0.52 | 1.71 | 0.62 | 31 |
| | | OC NAA | 8 | 1 | 9.44 | 0.13 | 9.07 | 0.59 | 10 | 0 | 12.35 | 0.70 | 12.38 | 0.58 | 11 | 0 | 15.76 | 2.13 | 15.46 | 1.54 | 29 |
| | | C-Reactive Protein | 9 | 0 | -0.11 | 0.12 | -0.05 | 0.44 | 10 | 0 | -0.06 | 0.50 | -0.11 | 0.60 | 15 | 0 | -0.29 | 0.47 | -0.19 | 0.67 | 34 |
| | | Interleukin 6 | 9 | 0 | -0.33 | 0.22 | -0.38 | 0.22 | 10 | 0 | -0.50 | 0.06 | -0.53 | 0.11 | 15 | 0 | -0.46 | 0.20 | -0.43 | 0.34 | 34 |
| | Patients | ACC Glutamate | 17 | 0 | 8.22 | 0.98 | 8.19 | 0.88 | 20 | 1 | 12.79 | 1.65 | 12.78 | 1.36 | 17 | 0 | 13.09 | 0.53 | 12.97 | 0.90 | 54 |
| | | ACC Glutamine | 17 | 0 | 4.35 | 0.67 | 4.39 | 1.08 | 19 | 1 | 4.30 | 0.50 | 4.11 | 1.26 | 17 | 0 | 3.50 | 0.59 | 3.57 | 0.51 | 53 |
| | | ACC Glutathione | 16 | 0 | 3.40 | 0.68 | 3.25 | 0.65 | 21 | 0 | 3.30 | 0.95 | 3.32 | 0.95 | 17 | 0 | 3.21 | 0.28 | 3.18 | 0.27 | 54 |
| | | ACC Creatine | 17 | 0 | 4.84 | 0.64 | 4.89 | 0.44 | 21 | 0 | 7.81 | 0.73 | 7.79 | 0.62 | 17 | 0 | 12.29 | 0.53 | 12.10 | 0.74 | 55 |
| | | ACC GABA | 12 | 2 | 2.19 | 0.80 | 2.17 | 0.60 | 21 | 0 | 2.31 | 0.24 | 2.21 | 0.39 | 16 | 0 | 0.62 | 0.30 | 0.59 | 0.24 | 49 |
| | | ACC NAA | 17 | 0 | 8.53 | 0.93 | 8.45 | 0.91 | 21 | 0 | 10.89 | 0.61 | 10.93 | 0.58 | 17 | 0 | 13.79 | 0.61 | 13.63 | 0.56 | 55 |
| | | OC Glutamate | 16 | 0 | 7.59 | 1.93 | 7.36 | 2.15 | 21 | 0 | 12.08 | 1.47 | 11.87 | 1.04 | 15 | 0 | 14.26 | 1.79 | 14.12 | 1.97 | 52 |
| | | OC Glutamine | 16 | 0 | 0.80 | 1.10 | 1.10 | 1.05 | 20 | 1 | 3.52 | 0.30 | 3.57 | 0.67 | 15 | 0 | 3.61 | 0.86 | 3.72 | 0.80 | 51 |
| | | OC Creatine | 16 | 0 | 5.64 | 0.36 | 5.83 | 0.94 | 21 | 0 | 8.42 | 0.53 | 8.40 | 0.50 | 15 | 0 | 13.28 | 2.18 | 13.08 | 2.06 | 52 |
| | | OC Glutathione | 16 | 0 | 2.56 | 0.65 | 2.56 | 0.50 | 21 | 0 | 3.62 | 0.62 | 3.67 | 0.94 | 15 | 0 | 2.85 | 0.33 | 2.83 | 0.50 | 52 |
| | | OC GABA | 14 | 0 | 2.70 | 0.30 | 2.71 | 0.28 | 21 | 0 | 2.81 | 0.21 | 2.80 | 0.30 | 13 | 0 | 1.49 | 0.44 | 1.63 | 0.55 | 48 |
| | | OC NAA | 16 | 0 | 9.06 | 1.72 | 9.19 | 2.02 | 21 | 0 | 12.22 | 0.37 | 12.26 | 0.53 | 15 | 0 | 15.93 | 1.87 | 15.76 | 2.11 | 52 |
| | | C-Reactive Protein | 16 | 0 | 0.20 | 0.45 | 0.09 | 0.52 | 20 | 0 | -0.18 | 0.77 | -0.11 | 0.73 | 21 | 0 | -0.09 | 0.74 | -0.12 | 0.49 | 57 |
| | | Interleukin 6 | 16 | 0 | -0.40 | 0.22 | -0.36 | 0.23 | 20 | 0 | -0.37 | 0.28 | -0.38 | 0.37 | 21 | 0 | -0.36 | 0.27 | -0.36 | 0.24 | 57 |

*Supplementary Table 4: Descriptive statistics for raw MRS and cytokine value at each site, pooled across participant groups.*

| Voxel | Metabolite | Cardiff | | | | | | Manchester | | | | | | Nottingham | | | | | | Total |
|---|---|---|---|---|---|---|---|---|---|---|---|---|---|---|---|---|---|---|---|---|
| | | N included | N extremes | Median | MAD | Mean | SD | N included | N extremes | Median | MAD | Mean | SD | N included | N extremes | Median | MAD | Mean | SD | N included |
| ACC | Glutamate | 58 | 0 | 7.68 | 1.52 | 7.45 | 1.59 | 64 | 2 | 12.14 | 1.65 | 12.47 | 1.95 | 61 | 1 | 13.09 | 0.93 | 12.98 | 0.99 | 183 |
| | Glutamine | 57 | 1 | 4.20 | 0.95 | 4.21 | 1.23 | 62 | 1 | 4.30 | 1.25 | 4.39 | 1.69 | 61 | 1 | 3.50 | 0.46 | 3.52 | 0.47 | 180 |
| | Glutathione | 56 | 0 | 2.94 | 0.86 | 3.01 | 0.77 | 64 | 0 | 2.99 | 0.93 | 3.04 | 0.92 | 61 | 1 | 3.27 | 0.39 | 3.26 | 0.39 | 181 |
| | GABA | 52 | 3 | 2.17 | 0.73 | 2.20 | 0.78 | 64 | 1 | 2.28 | 0.37 | 2.20 | 0.47 | 54 | 0 | 0.70 | 0.33 | 0.73 | 0.36 | 170 |
| | Creatine | 58 | 0 | 4.88 | 0.68 | 4.89 | 0.86 | 64 | 2 | 7.94 | 0.67 | 8.02 | 0.78 | 61 | 1 | 12.32 | 0.89 | 12.36 | 0.84 | 183 |
| | NAA | 58 | 0 | 8.36 | 1.26 | 8.21 | 1.35 | 66 | 0 | 10.92 | 0.65 | 10.95 | 0.70 | 60 | 2 | 13.91 | 0.65 | 13.86 | 0.79 | 184 |
| OC | Glutamate | 53 | 0 | 6.90 | 1.69 | 7.12 | 2.03 | 65 | 0 | 11.46 | 1.22 | 11.47 | 1.23 | 52 | 0 | 13.69 | 1.52 | 13.68 | 1.55 | 170 |
| | Glutamine | 53 | 0 | 1.01 | 1.22 | 1.23 | 1.31 | 63 | 1 | 3.72 | 0.83 | 3.82 | 1.04 | 51 | 1 | 3.55 | 0.53 | 3.65 | 0.67 | 167 |
| | Glutathione | 54 | 0 | 2.43 | 0.43 | 2.46 | 0.42 | 66 | 0 | 3.49 | 1.16 | 3.36 | 1.08 | 50 | 2 | 2.78 | 0.28 | 2.80 | 0.39 | 170 |
| | GABA | 56 | 0 | 2.74 | 0.37 | 2.75 | 0.33 | 66 | 0 | 2.77 | 0.27 | 2.75 | 0.33 | 49 | 0 | 1.53 | 0.53 | 1.65 | 0.54 | 171 |
| | Creatine | 52 | 1 | 5.92 | 0.81 | 6.19 | 0.98 | 65 | 0 | 8.63 | 0.71 | 8.66 | 0.61 | 52 | 0 | 13.34 | 1.36 | 13.31 | 1.64 | 169 |
| | NAA | 51 | 2 | 9.15 | 1.10 | 9.28 | 1.54 | 65 | 0 | 12.39 | 0.61 | 12.41 | 0.61 | 52 | 0 | 15.63 | 1.80 | 15.60 | 1.82 | 168 |

*Supplementary Table 5: Robust t statistics (tstat) and robust effect sizes (AKP, a robust version of Cohen's d) for effects of Diagnosis within each Phase group with bootstrapped 95% Confidence Intervals (95% CI).*

| Phase Group | Dependent variable | tstat | DF | p | ES (AKP) | 95% CI for ES |
|---|---|---|---|---|---|---|
| Recent | ACC Glutamate | 0.57 | 39.9 | .574 | 0.13 | -.29, .56 |
| | ACC Glutamine | 1.24 | 44.1 | .223 | -0.28 | -.78, .19 |
| | ACC Glutathione | 0.19 | 33.7 | .850 | -0.05 | -.53, .43 |
| | ACC GABA | 0.9 | 37.8 | .375 | -0.21 | -.70, .27 |
| | OC Glutamate | 1.13 | 31.8 | .268 | 0.29 | -.20, .77 |
| | OC Glutamine | 0.1 | 34.5 | .917 | 0.03 | -.51, .55 |
| | OC Glutathione | 0.3 | 35.9 | .768 | -0.07 | -.53, .41 |
| | OC GABA | 0.43 | 29.1 | .669 | -0.11 | -.58, .45 |
| | Mean GABA | 0.5 | 41 | .453 | -0.12 | -.71, .32 |
| | C-Reactive Protein | 0.73 | 50.6 | .466 | 0.16 | -.28, .60 |
| | Interleukin 6 | 1.44 | 55 | .157 | 0.29 | -.11, .69 |
| Established | ACC Glutamate | 2.62 | 39.4 | .012 | -0.59 | -1.12, -.06 |
| | ACC Glutamine | 0.28 | 52 | .779 | 0.06 | -.38, .45 |
| | ACC Glutathione | 1.78 | 35.6 | .084 | -0.42 | -.95, .07 |
| | ACC GABA | 1.18 | 50.8 | .242 | -0.24 | -.71, .15 |
| | OC Glutamate | 0.9 | 43.9 | .375 | 0.2 | -.28, .61 |
| | OC Glutamine | 0 | 34 | .998 | 0 | -.48, .47 |
| | OC Glutathione | 0.47 | 46 | .642 | 0.1 | -.35, .61 |
| | OC GABA | 3.1 | 55.7 | .003 | -0.6 | -1.14, -.19 |
| | Mean GABA | 3.44 | 59.5 | .001 | -0.64 | -1.17, -.23 |
| | C-Reactive Protein | 4.51 | 40.3 | >.001 | 0.99 | .40, 1.69 |
| | Interleukin 6 | 3.58 | 47.1 | >.001 | 0.74 | .30, 1.22 |

#### 7. Tabulated results for ANOVAs on MRS variables

Supplementary Table 6 shows the results for our key hypothesis tests, numbered 1 to 11.

- *Hypothesis test 1*: The hypothesis for which the study was designed to have 80% power (beta=.2) at an alpha of 0.05, namely, to find a Phase x Diagnosis effect for ACC glutamate. No adjustment was made to the *p* value for this hypothesis test
- *Hypothesis tests 2 to 6*: These five additional *a priori* hypotheses were FDR corrected, with the critical value for the least significant result set at *p*<.025
- *Hypothesis tests 7 to 9*: tests for interactions between Diagnosis and other factors, which would impact interpretation of the predicted main effect of Diagnosis:
  - *Hypothesis test 7*: Mean GABA: test for evidence of an Phase x Diagnosis interaction
  - *Hypothesis tests 8 & 9*: Within-subject Voxel differences in GABA: tests for within-between interactions between Voxel and Phase and/or Diagnosis, conducted by running t3way on within-subject voxel differences in site-normalised GABA values
- *Hypothesis tests 10 & 11:* unadjusted results for ACC glutamine.

Supplementary Table 7 gives group trimmed means (tr.mean) and standard errors (tr.SE) for each MRS variable of interest (trim= 20%).

*Supplementary Table 6: results for key hypothesis tests. Significant results (corrected as indicated) are shown in Bold with an **

| Hypothesis | Voxel | MRS | Effect | Q | N | p | Rank | Critical value | Adjusted p (q) |
|---|---|---|---|---|---|---|---|---|---|
| | **ACC Glutamate (alpha = .05, beta=.2)** | | | | | | | | |
| 1 | ACC | Glutamate | Phase x Diagnosis | 5.74 | 183 | **.022*** | | .050 | |
| | **Additional hypotheses with FDR correction** | | | | | | | | |
| 2 | ACC | Glutathione | Phase x Diagnosis | 1.89 | 181 | .175 | 3 | .025 | .350 |
| 3 | ACC | Glutathione | Diagnosis | 1.70 | 181 | .197 | 4 | .033 | .296 |
| 4 | ACC | GABA | Diagnosis | 1.07 | 170 | .306 | 5 | .042 | .367 |
| 5 | OC | GABA | Diagnosis | 4.99 | 171 | .030 | 2 | .017 | .090 |
| 6 | Voxel mean | GABA | Diagnosis | 8.08 | 155 | .006 | 1 | .008 | **.036*** |
| | **GABA: tests for interactions** | | | | | | | | |
| 7 | Mean | GABA | Phase x Diagnosis | 2.76 | 155 | .102 | | | Unc. |
| 8 | Difference | GABA | Phase x Diagnosis | .07 | 155 | .794 | | | Unc. |
| 9 | Difference | GABA | Phase | .29 | 155 | .600 | | | Unc. |
| | **ACC Glutamine** | | | | | | | | |
| 10 | ACC | Glutamine | Phase x Diagnosis | .60 | 180 | .440 | | | Unc. |
| 11 | ACC | Glutamine | Diagnosis | .16 | 180 | .694 | | | Unc. |

*Supplementary Table 7: Group trimmed means (tr.mean) and trimmed standard errors (tr.SE) for each MRS variable of interest (trim= 20%).*

| Voxel | MRS | Phase group | Controls tr.mean (Tr.SE) | Patients tr.mean (Tr.SE) |
|---|---|---|---|---|
| ACC | Glutamate | Recent | 0.14 (0.16) | 0.24 (0.11) |
| | | Established | 0.23 (0.21) | -0.42 (0.15) |
| | Glutamine | Recent | 0.20 (0.17) | -0.04 (0.13) |
| | | Established | -0.17 (0.18) | -0.09 (0.19) |
| | Glutathione | Recent | 0.19 (0.20) | 0.15 (0.12) |
| | | Established | 0.11 (0.20) | -0.30 (0.13) |
| | GABA | Recent | 0.08 (0.18) | -0.12 (0.14) |
| | | Established | 0.12 (0.17) | -0.16 (0.17) |
| OC | GABA | Recent | 0.22 (0.21) | 0.12 (0.12) |
| | | Established | 0.34 (0.13) | -0.29 (0.17) |
| Mean | GABA | Recent | 0.16 (0.17) | 0.02 (0.08) |
| | | Established | 0.27 (0.09) | -0.30 (0.14) |

### 8. Correlation analysis (see main fig 2)

*Supplementary Figure 1: Scattergrams and distributions for pairwise correlations between MRS, cytokine and clinical variables depicted in Figure 2 (main text) and Supplementary Table 8 below.*

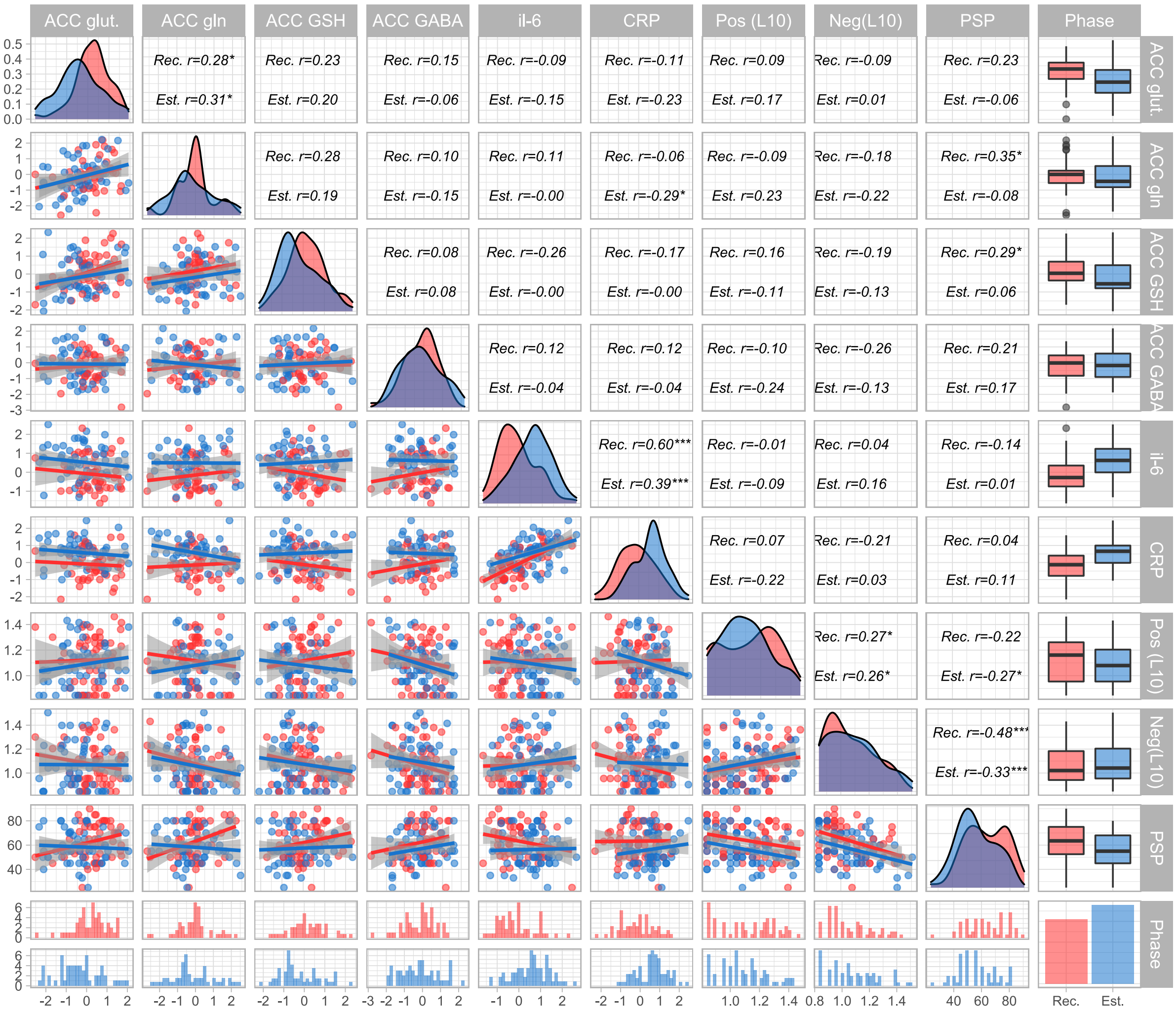


Red=Recent; Blue=Established. **p*<.05; ***p*<.01; ***p<.001; † correlations differ between phase groups at *p*<.05.

*Supplementary Table 8: Correlations for all patients between MRS variables and potential influences (age, drug exposure, BMI and cytokines), and clinical measures (positive and negative symptoms, IQ and PSP), after group-mean-centering. *=p<.05 unc.; **-p<.01, unc.; ***-p<.001 unc. † Correlations where within-phase correlations differed at p<.05.*

**Correlations across all patients**

*(Minimum N=97, all variables group-mean centered)*

| | ACC glu | ACC gln | ACC GSH | ACC GABA | OC GABA | age | AP day | AP life | Ill Dur | BMI | il 6 | crp | Pos | Neg | IQ |
|---|---|---|---|---|---|---|---|---|---|---|---|---|---|---|---|
| **ACC gln** | .36*** | | | | | | | | | | | | | | |
| **ACC GSH** | .21* | .18 | | | | | | | | | | | | | |
| **ACC GABA** | .01 | .00 | .06 | | | | | | | | | | | | |
| **OC GABA** | -.13 | -.02 | .18 | .21* | | | | | | | | | | | |
| **age** | .06 | -.07 | -.31*** | .09 | .01 | | | | | | | | | | |
| **AP-day** | -.15 | -.17 | .01 | -.03 | -.08 | -.01 | | | | | | | | | |
| **AP-life** | -.01 | -.25** | .03 | .00 | -.08 | .15 | .35† | | | | | | | | |
| **Ill Dur** | .10 | -.23* | -.24† | .00 | -.15 | .51† | .02 | .24† | | | | | | | |
| **BMI** | -.12 | -.22* | -.12 | .00 | .12 | .06 | .12† | .24** | .04 | | | | | | |
| **il-6** | -.16 | .08 | -.11† | .02 | .09 | .04 | .04 | -.20* | -.11 | .26** | | | | | |
| **crp** | -.19* | -.12 | -.08 | .06 | .18 | .17 | .10 | .05 | .05 | .29*** | .50† | | | | |
| **Pos** | .14 | .07 | .04 | -.24* | .00 | .12 | .03 | -.08 | .06 | -.08 | -.06 | -.10 | | | |
| **Neg** | -.05 | -.14 | -.15 | -.20* | -.17 | -.06 | .10 | -.06† | -.02 | -.02 | .11 | -.07 | .26** | | |
| **IQ** | .03 | .01 | .14 | .11† | -.16 | .03 | -.04 | .04 | .06 | -.16 | -.17 | -.27** | .05 | -.18* | |
| **psp** | .07 | .13† | .18 | .19 | .05 | -.10 | .03 | -.08 | .08 | -.04 | -.06 | .06 | -.24** | -.39*** | .15 |

**Correlations within Recent group**

*Minimum N=47*

| | ACC glu | ACC gln | ACC GSH | ACC GABA | OC GABA | age | AP day | AP life | Ill Dur | BMI | il 6 | crp | Pos | Neg | IQ |
|---|---|---|---|---|---|---|---|---|---|---|---|---|---|---|---|
| **ACC gln** | .29* | | | | | | | | | | | | | | |
| **ACC GSH** | .23 | .24 | | | | | | | | | | | | | |
| **ACC GABA** | .15 | .06 | .08 | | | | | | | | | | | | |
| **OC GABA** | -.20 | -.08 | .06 | .18 | | | | | | | | | | | |
| **age** | -.10 | .05 | -.22 | .11 | .10 | | | | | | | | | | |
| **AP-day** | .02 | -.16 | .12 | .04 | .05 | .05 | | | | | | | | | |
| **AP-life** | .12 | -.23 | .18 | -.04 | -.05 | .01 | .65*** | | | | | | | | |
| **Ill Dur** | .01 | -.08 | .08 | .12 | -.03 | -.02 | -.24 | -.20 | | | | | | | |
| **BMI** | .04 | -.16 | -.19 | -.05 | .02 | .21 | .36** | .33* | -.01 | | | | | | |
| **il-6** | -.14 | .13 | -.30* | .11 | -.01 | .18 | .00 | -.27* | .11 | .21 | | | | | |
| **crp** | -.13 | -.01 | -.17 | .12 | .15 | .32* | .23 | -.04 | .07 | .24 | .64*** | | | | |
| **Pos** | .09 | -.09 | .16 | -.10 | -.04 | .01 | .06 | -.02 | .19 | -.01 | -.03 | .04 | | | |
| **Neg** | -.09 | -.18 | -.19 | -.26 | -.22 | .00 | .02 | .17 | .03 | .10 | .06 | -.19 | .27* | | |
| **IQ** | .04 | -.11 | .17 | .32* | -.16 | -.05 | -.13 | -.02 | .31* | -.17 | -.31* | -.27* | .01 | -.14 | |
| **psp** | .23 | .29* | .29* | .21 | -.01 | -.25 | .04 | -.01 | -.11 | -.12 | -.13 | .00 | -.22 | -.48*** | .19 |

**Correlations within Established group**

*Minimum N=52*

| | ACC glu | ACC gln | ACC GSH | ACC GABA | OC GABA | age | AP day | AP life | Ill Dur | BMI | il 6 | crp | Pos | Neg | IQ |
|---|---|---|---|---|---|---|---|---|---|---|---|---|---|---|---|
| **ACC gln** | .41** | | | | | | | | | | | | | | |
| **ACC GSH** | .19 | .17 | | | | | | | | | | | | | |
| **ACC GABA** | -.05 | .01 | .05 | | | | | | | | | | | | |
| **OC GABA** | -.08 | .01 | .28* | .22 | | | | | | | | | | | |
| **age** | .14 | -.12 | -.35** | .06 | -.02 | | | | | | | | | | |
| **AP-day** | -.24 | -.17 | -.05 | -.08 | -.13 | -.04 | | | | | | | | | |
| **AP-life** | -.05 | -.26* | -.05 | .01 | -.09 | .18 | .24* | | | | | | | | |
| **Ill Dur** | .11 | -.26* | -.32* | .02 | -.15 | .68*** | .04 | .33** | | | | | | | |
| **BMI** | -.22 | -.25* | -.07 | .04 | .16 | -.02 | .00 | .19 | -.02 | | | | | | |
| **il-6** | -.20 | .00 | .05 | -.03 | .14 | -.04 | .02 | -.18 | -.15 | .32** | | | | | |
| **crp** | -.23 | -.23 | .01 | .00 | .20 | .09 | -.02 | .08 | .06 | .33** | .39*** | | | | |
| **Pos** | .21 | .22 | -.07 | -.35** | .03 | .22 | .01 | -.14 | .05 | -.12 | -.09 | -.22 | | | |
| **Neg** | -.01 | -.07 | -.12 | -.14 | -.15 | -.09 | .14 | -.19 | -.04 | -.10 | .16 | .03 | .26* | | |
| **IQ** | .03 | .10 | .12 | -.02 | -.13 | .08 | .04 | .09 | .03 | -.15 | -.07 | -.27* | .08 | -.21 | |
| **psp** | -.06 | -.03 | .05 | .17 | .11 | .01 | .04 | -.12 | .13 | .00 | .01 | .11 | -.27* | -.33** | .11 |

*Supplementary Figure 2: Scatterplots, distributions and robust correlations within and between confounders (AP-day, AP-life, BMI), mechanistic mediators (cytokines) and ACC glutamate and glutamine expressed as difference from age-matched control mean (ACC gluΔ, ACC gln Δ).*

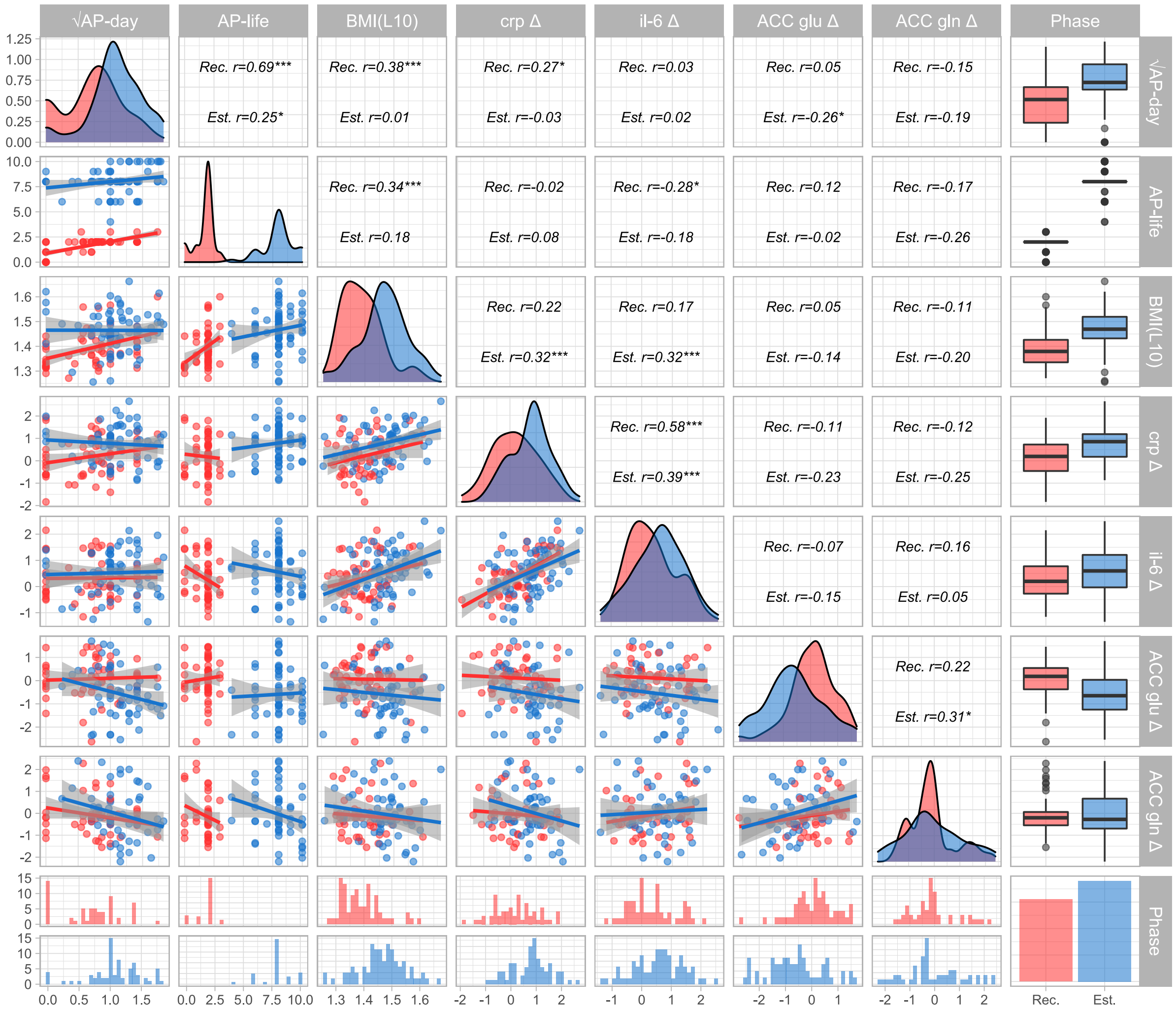


Red=Recent; Blue=Established. *$p$<.05; **$p$<.01; ***p<.001; † correlations differ between phase groups at $p$<.05.

#### 9. Mediation analysis

The confirmation of our primary prediction of a greater ACC glutamate deficit in Established than in Recent onset patients was consistent with our neurotoxic model. However, as both AP-day and AP-life were substantially higher in the Established group, it would also be consistent with evidence that reduced glutamate in older patients is associated with greater medication exposure [3]. We therefore evaluated the evidence for each of these explanations through a series of mediation models.

Phase was the primary predictor in all models, with the Recent group coded as the reference category. Positive standardised parameters (denoted *β*) for direct effects of Phase therefore indicate higher predicted values in the Established group, and negative parameters indicate lower predicted values.

We first expressed relevant patient variables (ACC glutamate; ACC glutamine, CRP; IL-6) in terms of deviation from the expected value for their age-group by subtracting the mean value of their age-matched control-group from each patient's value. Age-group adjusted values are denoted by the suffix Δ. Supplementary Figure 2 shows pairwise relationships between these variables and with three potential medication-related confounders, AP-day, AP-life (square root transformed) and BMI ($log_{10}$ transformed).

##### *Mediation of phase effects on ACC glutamate by antipsychotic medication.*

In the model shown in Supplementary Figure 3A, we found no evidence to suggest that the effect of Phase on ACC glutamate Δ (ACC glu Δ) was mediated by AP-day, *β* =-0.04, *p*=.240), while a negative direct effect of Phase remained evident, *β* =-0.29, *p*=.002. When we included AP-life as a second mediator (Figure 3A, main text), the negative direct effect of Phase remained evident, *β*=-0.57, *p*=.023, again with no evidence to indicate mediation by antipsychotic medication.

##### *Possible effects of antipsychotic medication on ACC glutamate via glutamine.*

ACC glutamine, was strongly positively correlated with ACC glutamate (*r*=.36, *p*<.001) across both phase groups, and also negatively correlated with AP-life (Supplementary Figure 2 above). Nonetheless, mean ACC gln Δ was not significantly lower in the Established Group, despite their greater AP exposure, suggestion possible suppression by AP-life of an underlying elevating effect of established illness on ACC gln Δ. An exploratory regression model with both Phase and AP-life as predictors of ACC gln Δ provided confirmatory evidence of higher ACC gln Δ in Established than in Recent patients, *β*=0.63, *p*=.016 when its negative association with AP-life, *β*=-0.65, *p*=.018 is controlled for.

We therefore investigated whether ACC gln Δ might mediate effects of Phase on ACC glu Δ, either via AP-life, or via an underlying effect of established illness on ACC gln Δ. This model (Supplementary Figuere 3B) provided weak evidence (*p*<.1) for partial mediation of the Phase effect on ACC Glutamate via the negative effect of AP-life on ACC glu Δ , *β*=-0.18, *p*=.066. However this mediation pathway was countered by a second indirect effect via the positive effect of Phase on ACC gln Δ, *β*=0.19, *p*=.059. Accounting for these indirect pathways strengthened the evidence for a direct effect of Phase on ACC glu Δ, *β*=-0.66, *p*=.002.

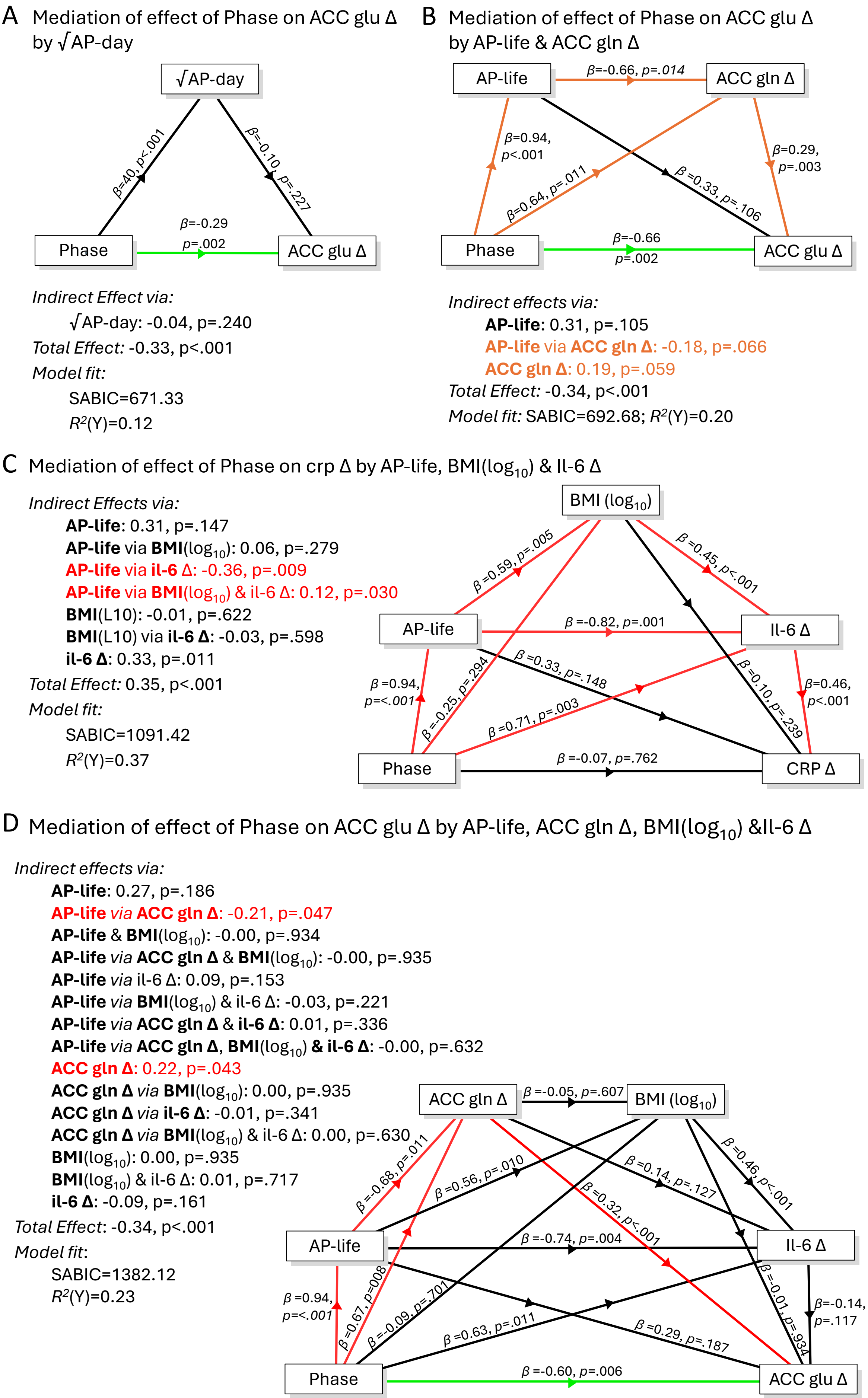

A Mediation of effect of Phase on ACC glu Δ by √AP-day
√AP-day
β=.40, p<.001
β=-0.10, p=.227
Phase
β=-0.29 p=.002
ACC glu Δ
Indirect Effect via:
√AP-day: -0.04, p=.240
Total Effect: -0.33, p<.001
Model fit:
SABIC=671.33
R²(Y)=0.12
B Mediation of effect of Phase on ACC glu Δ by AP-life & ACC gln Δ
AP-life
β=-0.66, p=.014
ACC gln Δ
β=0.94, p<.001
β=0.64, p=.011
β=0.33, p=.106
β=0.29, p=.003
Phase
β=-0.66 p=.002
ACC glu Δ
Indirect effects via:
AP-life: 0.31, p=.105
AP-life via ACC gln Δ: -0.18, p=.066
ACC gln Δ: 0.19, p=.059
Total Effect: -0.34, p<.001
Model fit: SABIC=692.68; R²(Y)=0.20
C Mediation of effect of Phase on crp Δ by AP-life, BMI(log10) & Il-6 Δ
Indirect Effects via:
AP-life: 0.31, p=.147
AP-life via BMI(log10): 0.06, p=.279
AP-life via il-6 Δ: -0.36, p=.009
AP-life via BMI(log10) & il-6 Δ: 0.12, p=.030
BMI(L10): -0.01, p=.622
BMI(L10) via il-6 Δ: -0.03, p=.598
il-6 Δ: 0.33, p=.011
Total Effect: 0.35, p<.001
Model fit:
SABIC=1091.42
R²(Y)=0.37
BMI (log10)
β=0.59, p=.005
β=0.45, p<.001
AP-life
β=-0.82, p=.001
Il-6 Δ
β=0.33, p=.148
β=0.10, p=.239
β=0.94, p=<.001
β=-0.25, p=.294
β=0.71, p=.003
β=0.46, p<.001
Phase
β=-0.07, p=.762
CRP Δ
D Mediation of effect of Phase on ACC glu Δ by AP-life, ACC gln Δ, BMI(log10) &Il-6 Δ
Indirect effects via:
AP-life: 0.27, p=.186
AP-life via ACC gln Δ: -0.21, p=.047
AP-life & BMI(log10): -0.00, p=.934
AP-life via ACC gln Δ & BMI(log10): -0.00, p=.935
AP-life via il-6 Δ: 0.09, p=.153
AP-life via BMI(log10) & il-6 Δ: -0.03, p=.221
AP-life via ACC gln Δ & il-6 Δ: 0.01, p=.336
AP-life via ACC gln Δ, BMI(log10) & il-6 Δ: -0.00, p=.632
ACC gln Δ: 0.22, p=.043
ACC gln Δ via BMI(log10): 0.00, p=.935
ACC gln Δ via il-6 Δ: -0.01, p=.341
ACC gln Δ via BMI(log10) & il-6 Δ: 0.00, p=.630
BMI(log10): 0.00, p=.935
BMI(log10) & il-6 Δ: 0.01, p=.717
il-6 Δ: -0.09, p=.161
Total Effect: -0.34, p<.001
Model fit:
SABIC=1382.12
R²(Y)=0.23
ACC gln Δ
β=-0.05, p=.607
BMI (log10)
β=-0.68, p=.011
β=0.56, p=.010
β=0.14, p=.127
β=0.46, p<.001
β=0.32, p<.001
AP-life
β=-0.74, p=.004
Il-6 Δ
β=0.94, p=<.001
β=0.67, p=.008
β=-0.09, p=.701
β=0.63, p=.011
β=0.29, p=.187
β=-0.01, p=.934
β=-0.14, p=.117
Phase
β=-0.60, p=.006
ACC glu Δ

*Supplementary Figure 3 Panel A: There was no evidence that higher AP-day in the Established group mediated the greater ACC glutamate deficit in the Established group. Panel B: there was weak evidence (p<.1) for partial mediation of the Phase effect on ACC glu Δ by ACC gln via two pathways, shown in orange), with opposite signs. Panel C: The Phase effect on CRP (greater elevation relative to controls in the Established than in the Recent group) was fully mediated via three pathways all of which included Il-6, two of which were associated with the effects of AP-life on Il-6. Panel D: Phase effects on ACC glu Δ*

#### *Possible effects of antipsychotic medication on ACC glutamate via cytokines.*

ACC glutamate tended to be negatively correlated with the two inflammatory cytokines of interest, CRP and Il-6, suggesting a possible shared underlying mechanism. CRP and Il-6 were also strongly correlated both with each other and with BMI, and BMI, in turn, showed the expected strong positive association with AP-life. However, despite this association, greater AP-life was associated with lower il-6 across both phase groups.

In the model shown in Figure 3B (main text), we investigated to what extent the greater CRP Δ levels in the established group were mediated by AP-life, either directly or via its effects on BMI, and found that phase-related elevation of CRP-Δ was fully accounted for by the effect of AP-life on BMI. We then investigated whether this effect might itself be mediated by effects of BMI on IL-6 Δ (Supplementary Figure 3C). This model indicated that the effects of Phase on CRP Δ was mediated via three separate indirect pathways, all involving il-6 Δ:

- A net positive effect of Phase on il-6 Δ via the effects of AP-life on BMI
- A net negative effect of Phase via the negative effects of AP-life on il-6 Δ
- A net positive effect of Phase on il-6 Δ

These effects Phase on il-6 Δ are shown in Figure 3C (main text) and indicated a "two-handed" effect of AP-life on il-6-Δ: a positive effect via BMI that only partly counteracts a more direct negative effect. Moreover, this "two-handed" effect of AP-life suppresses the effects of an underlying residual Phase-related il-6 Δ excess in the Established group, which emerges when both medication effects are controlled for.

Finally, we investigated whether this apparently Phase-related excess il-6 Δ might be related to the Phase-related reduction in ACC glu Δ, possibly via a shared Phase-related inflammatory process. In the model shown in Supplementary Figure 3D, we included AP-life, ACC gln Δ, BMI and Il-6 Δ as serial mediators, to control for any effects of AP-life mediated via ACC gln Δ, or BMI. A simplified version of this model, in which we treated AP-life, ACC gln Δ, and BMI as covariates is shown in Figure 3D (main text). Neither of these models provided evidence that the ACC glutamate deficit in Established patients was mediated by phase-related excess il-6, and in both models the effect of Phase on ACC glu Δ remained evident. The four-mediator model slightly strengthened the evidence that the negative effect of AP-life on ACC gln Δ partially mediates the effect of Established illness in reducing ACC glu Δ, but also strengthened the evidence for an association between the underlying ACC glutamate deficit in Established patients and higher levels of ACC glutamine than predicted by their greater antipsychotic exposure.
